# Lying in wait: the resurgence of dengue virus after the Zika epidemic in Brazil

**DOI:** 10.1101/2020.08.10.20172247

**Authors:** Anderson Fernandes Brito, Lais Ceschini Machado, Márcio Junio Lima Siconelli, Rachel J. Oidtman, Joseph R. Fauver, Rodrigo Dias de Oliveira Carvalho, Filipe Zimmer Dezordi, Mylena Ribeiro Pereira, Luiza Antunes de Castro-Jorge, Elaine Cristina Manini Minto, Luzia Márcia Romanholi Passos, Chaney C. Kalinich, Mary E. Petrone, Emma Allen, Guido Camargo España, Angkana T. Huang, Derek A. T. Cummings, Guy Baele, Rafael Freitas Oliveira Franca, T. Alex Perkins, Benedito Antônio Lopes da Fonseca, Gabriel Luz Wallau, Nathan D. Grubaugh

## Abstract

After Zika virus (ZIKV) emerged and caused an epidemic in the Americas in 2016, both Zika and dengue incidence declined in the following years (2017-2018) to a record low in many countries. Following this period of low incidence, dengue resurged in 2019 in Brazil, causing ~2.1 million cases. The reasons for the recent fluctuations in dengue incidence and the maintenance of dengue virus (DENV) through periods of low transmission are unknown. To investigate this, we used a combination of epidemiological and climatological data to estimate dengue force of infection (FOI) and model mosquito-borne transmission suitability since the early 2000s in Brazil. Our estimates of FOI revealed that the rate of DENV transmission in 2018-2019 was exceptionally low, due to a low proportion of susceptible population rather than changes to ecological conditions. This supports the hypothesis that the synchronous decline of dengue in Brazil may be explained by protective immunity from pre-exposure to ZIKV and/or DENV in prior years. Furthermore, we sequenced 69 genomes of dengue virus serotype 1 (DENV-1) and DENV-2 circulating in Northeast and Southeast Brazil, and performed phylogeographic analyses to uncover patterns of viral spread. We found that the outbreaks in Brazil in 2019 were caused by DENV lineages that were circulating locally prior to the Zika epidemic and spread cryptically during the period of low transmission. Despite the period of low transmission, endemic DENV lineages persisted for 5-10 years in Brazil before causing major outbreaks. Our study challenges the paradigm that dengue outbreaks are caused by recently introduced new lineages, but rather they may be driven by established lineages circulating at low levels until the conditions are conducive for outbreaks.

## Introduction

After Zika virus (ZIKV) was introduced in Brazil around late 2013^12^, the resulting epidemic reached its peak in March 2016. The decline of Zika cases in Brazil during 2017 and 2018 was mirrored by a decline in reported dengue cases. Interestingly, similar patterns were observed throughout the Americas^3^. The two years of abnormally low dengue incidence were followed by a synchronized resurgence of cases in 2019 across the Americas. That year a record high 3.1 million dengue cases were reported throughout the region (2.1 million in Brazil alone), with an incidence of 321.58 cases per 100,000 habitants^4^. These fluctuations in dengue incidence raise several important questions: (***1***) Did DENV transmission fluctuate or just reported symptomatic dengue cases? (***2***) Can Zika outbreaks impact subsequent dengue virus (DENV) transmission? (***3***) Was the 2019 resurgence in dengue caused by new DENV introductions? Considering the global distribution of both ZIKV and DENV^5,6^, answering these questions can help us to better prepare for and control future outbreaks.

Several anthropological, epidemiological, and ecological factors may explain the decline of dengue incidence post Zika outbreaks. Human interventions, such as enhanced mosquito control, or changes in human behaviour in response to the Zika epidemic could explain the decline in reported dengue cases^7,8^. However, these changes driven by human intervention would not be expected to simultaneously occur across a diverse region. Ecological factors, such as local humidity and temperature may also impact the transmission dynamics of arboviral diseases^9^. As the mosquito vectors for DENV, *Aedes aegypti* and *Ae. albopictus* can transmit ZIKV and chikungunya virus (CHIKV), changes in climate would alter the transmission of all three viruses^7-9^, and could be at least partially responsible for the fluctuations in DENV. Finally, immunological cross-protection of flavivirus antibodies can also impact epidemics^10^, and the potential effects of a previous exposure to ZIKV on secondary DENV infections have been debated. Previous studies suggest that pre-exposure to ZIKV could (***1***) lead to severe dengue disease due to antibody-dependent enhancement^11^, as observed in secondary dengue infections by distinct dengue serotypes^12,13^; and/or (***2***) provide partial protection, leading to less severe disease^8,14^. With nearly two-thirds of the dengue cases being mild or asymptomatic^6^, high levels of cross-protection could lead to less severe infections, and even higher underreporting. This last scenario could explain the declines in dengue cases observed in countries hard hit by ZIKV prior to dengue resurgence in 2019, like in Brazil^15^, Bolivia^16^, Suriname^17^, and Nicaragua^18^.

The resurgence of dengue raises questions about the origin of the 2019 outbreaks in Brazil. All four DENV serotypes have circulated throughout the Americas since their reintroductions into the western hemisphere in the 1970-80s^19-23^; however, in more recent years, DENV serotype 1 (DENV-1) and DENV-2 have become predominant in Brazil^24^. Thus, it is unclear if the recent outbreaks were caused by existing DENV lineages cryptically circulating in Brazil before and during the years of low reported dengue cases, or if they were the results of new DENV lineages introduced from other countries. Historical dengue outbreaks caused by DENV-2 in Puerto Rico, for example, have been caused by both reintroductions (clade replacement) and local survival through periods of low transmission^25^. The distinction is important for both our basic understanding of DENV maintenance and developing targeted control methods.

To answer these questions, we combined genomic, epidemiological, and ecological data to investigate the resurgence of dengue across Brazil and within two distinct geographic regions. Our estimates of DENV transmission show that the periods of low incidence in 2017-2018 were caused by low population susceptibility rather than environmental conditions or surveillance gaps. We then sequenced DENV-1 and DENV-2 from 69 clinical samples collected during 2010, 2018, and 2019 from Northeast and Southeast Brazil to identify the origins and trace the spread of the viruses during the resurgence in 2019. We found that the recent dengue outbreaks were caused by DENV lineages circulating in Brazil before the Zika epidemic, meaning that the lineages endured the period of low transmission. Furthermore, the dispersal velocity of these persistent DENV lineages peaked in 2016 and slowed with the decline of Zika and dengue in 2017. Together, our results indicate that DENV established cryptic transmission for more than five years before causing outbreaks in 2019, perhaps due to waning cross-protection provided by existing DENV and/or ZIKV population immunity^14,26^, a scenario that may have occurred across the Americas.

## Results

### Patterns of dengue incidence after the Zika epidemic

While the annual dengue incidence in Brazil varies by year, the general trend in the 21st century has been an increasing burden of disease (**Figure 1A**). In 2016, dengue cases exceeded 1.6 million, which was a record at the time^27^. In 2017 and 2018, the number of annually reported dengue cases declined to ~250,000, the lowest since 2005 (~200,000; **Figure 1A**). In 2019, dengue cases again rebounded, setting a new record of ~2.1 million reported cases in Brazil (**Figure 1A**). We show that similar patterns of dengue incidence were also observed in each of the five geographic regions of the country (**Figure 1B-C**).

From 2016 to 2018, Zika incidence followed a similar pattern as dengue. After Zika was introduced in the country in late 2013^12^, and spread undetected until May 2015^28^, the number of Zika cases peaked in March 2016 (**Figure 1A, Figure S1**). Subsequently, dengue cases dropped drastically across the country, only to resurge in 2019. One hypothesis for the sudden decrease in dengue incidence during 2017 and 2018 is that ZIKV infections provided some cross-protective immunity against DENV infections^8,14^. If accurate, the rise in dengue incidence during 2019 could represent waning cross-protection.

**Figure 1.**
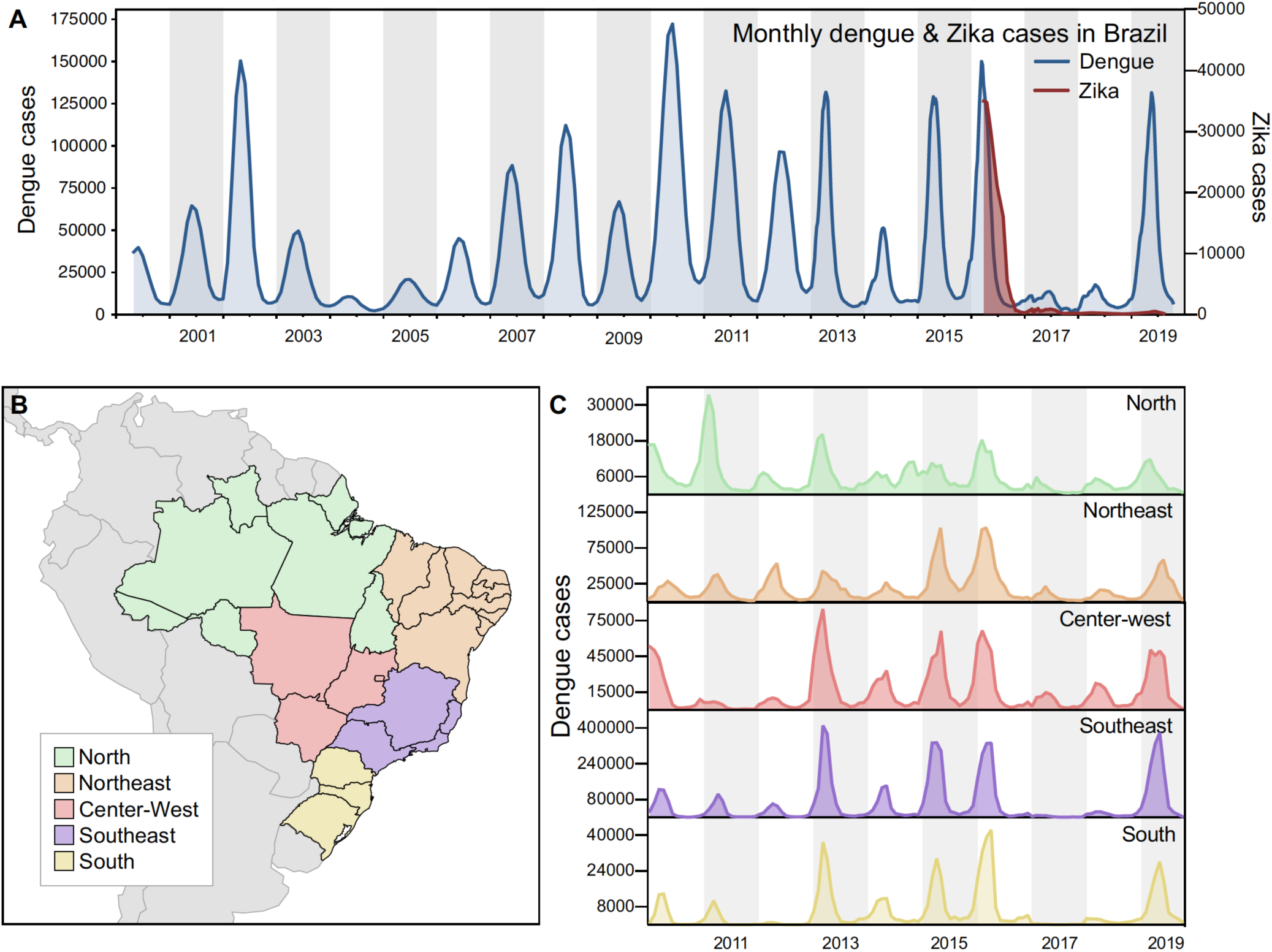
Dengue resurgence in Brazil following a 2-year drought post Zika epidemic. (**A**) Monthly dengue and Zika cases in Brazil, from 2000 to 2019. (**B**) Map of geographic regions in Brazil. (**C**) Dengue cases per geographic region in Brazil, from 2010 to 2019 (y-axis not in the same scale).

### Annual dengue virus force of infection

The trends in dengue incidence could be caused by DENV transmission dynamics and/or biases in surveillance systems (**Figure 1**). Commonly, only symptomatic infections are detected by surveillance, and the probability of experiencing dengue fever or severe dengue depends on whether an individual is experiencing a primary, secondary, or post-secondary DENV infection^29^. Changes in the demography and recent history of DENV transmission in a population can, through this mechanism, generate substantial inter-annual variability in the probability that individuals experience dengue fever or severe dengue in a given year^30-32^. To determine whether natural variation in these factors could explain the decline in dengue incidence in 2017 and 2018, we estimated the force of infection (FOI) of DENV transmission on an annual basis from 2002 through 2019. This metric, defined as the rate at which susceptible individuals become infected, accounts for the aforementioned natural changes in the probability of experiencing dengue fever or severe dengue and provides a more direct measure of DENV transmission than reported incidence.

Our estimates of FOI indicate that transmission was indeed low in 2017 and 2018 (**Figure 2**). When we ranked years by their FOI across the 100 replicates that comprised our estimates, we found that 2017 and 2018 were nearly always in the lower half of years in all five regions, but the lowest only in the Center-West region in2018 (**Figure S2**). Our inferred changes in the probability of dengue fever or severe dengue conditional on infection were flat from 2002 through 2019 (**Figure S3**), suggesting that changes in DENV transmission, not the a decline in secondary and post-secondary infections that would be more likely to be severe and to be captured by surveillance, likely contributed to the drop in dengue incidence in 2017 and 2018. Thus, our estimates of DENV transmission match the patterns in dengue incidence (**Figure 1**), supporting the scenario that dengue did decline following the Zika epidemic.

One contributing factor to the relatively low FOI in those years may have been that susceptibility to DENV infection appeared to be at or near its lowest in 2017 and 2018 (**Figure S4**). Relative to its maximum across 2002-2019, population susceptibility in 2017 and 2018 was around 5% lower in absolute terms and 7-10% lower in relative terms. This reduction in susceptibility in 2017 and 2018 was likely a result of high DENV transmission over several preceding years, which may have provided some within and between DENV serotype protection (though we do not have sufficient data on serotypes to analyze). Based on this analysis, however, we cannot exclude a possible role for other factors, such as cross-protection conferred by ZIKV infection^14,26^, as also contributing to a reduction in dengue incidence in those years.

**Figure 2.**
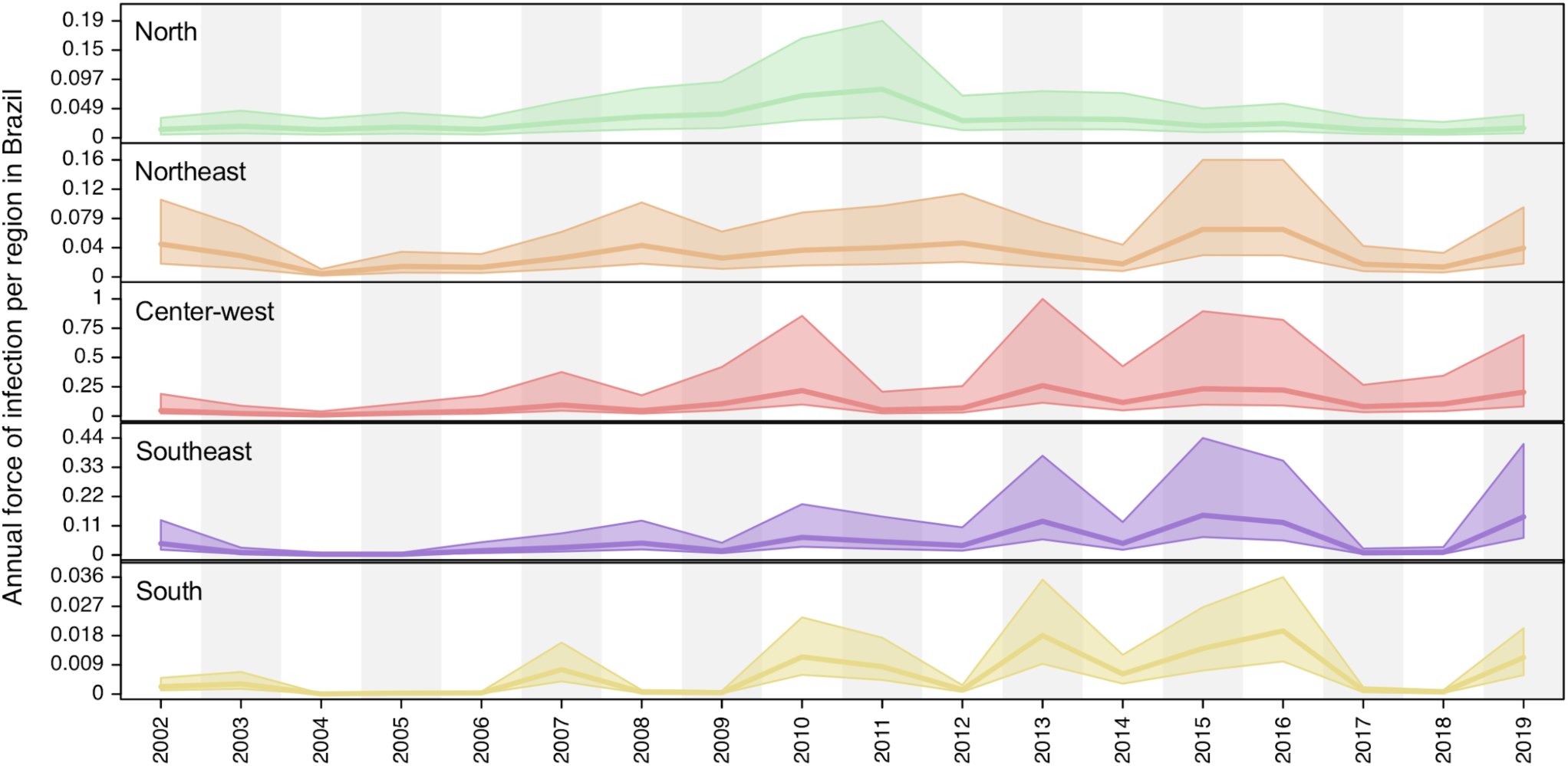
Dengue virus force of infection was low following the Zika epidemic. Shaded regions indicate the 95% credible intervals and dark lines indicate median estimate. The regions and colors correspond to the map in **Figure 1B**.

### Mosquito-borne virus surveillance and transmission potential

If past DENV and/or ZIKV infections provided immunological protection against DENV, then the same patterns of dengue decline should follow throughout the Americas in the wake of the Zika epidemic. We show that the drop in dengue incidence in 2017-2018 was also observed in many other regions in Latin America and the Caribbean, demonstrating this pattern was not unique to Brazil (**Figure 3A**). However, there could be other factors contributing to the patterns of dengue incidence that are unrelated to population immunity or susceptibility, such as: (***1***) changes in mosquito-borne virus surveillance practices, including surveillance “fatigue” (e.g. decrease in allocated resources) following an epidemic; and (***2***) changing ecological factors that govern mosquito-borne transmission.

To evaluate the potential impacts of mosquito-borne virus surveillance during and after the Zika epidemic, we investigated the patterns of reported cases of chikungunya (**Figure 3B**). CHIKV is also transmitted by *Ae. aegypti* and *Ae. albopictus*, but as an alphavirus, it would not be affected by previous exposure to the flavirususes, ZIKV and DENV. Thus, if CHIKV followed similar patterns as ZIKV and DENV, it would suggest that systematic changes - ecology and/or surveillance - would have also played a large role. We found that chikungunya cases, which also peaked in Brazil during 2016, gradually declined through 2019, and did not match the recent patterns of dengue incidence (**Figure 3B**). As the dengue patterns in Brazil match those found throughout the Americas (**Figure 3A**), but reported chikungunya cases in Brazil do not follow the same steep decline and rebound (**Figure 3B**), this may suggest that changes in surveillance are not a primary cause of the observed dengue patterns^8^ (**Figure 1**).

As dengue outbreaks within a region can be synchronous, driven in part by large climatic shifts (e.g. El Niño)^33-36^, we sought to determine if recent ecological changes could have altered virus transmission potential by *Ae. aegypti* mosquitoes. For this analysis, we used *Index P*, a metric that uses dynamic temperature and humidity data with epidemiological factors (**Figure 3C**)^37^, to estimate the transmission potential of mosquitoes from major urban areas in the regions historically most affected by dengue: Northeast and Southeast Brazil (**Figure 1B-C**). Our analysis shows that *Index P* was not notably different between 2016-2019 (**Figure 3C**), and does not account for the differences in dengue cases in those regions (**Figure 1C**). Thus, based on dengue incidence in the Americas, chikungunya incidence in Brazil, and *Index P*, we conclude that the changes in dengue patterns were likely not due to changing surveillance efforts or climate.

**Figure 3.**
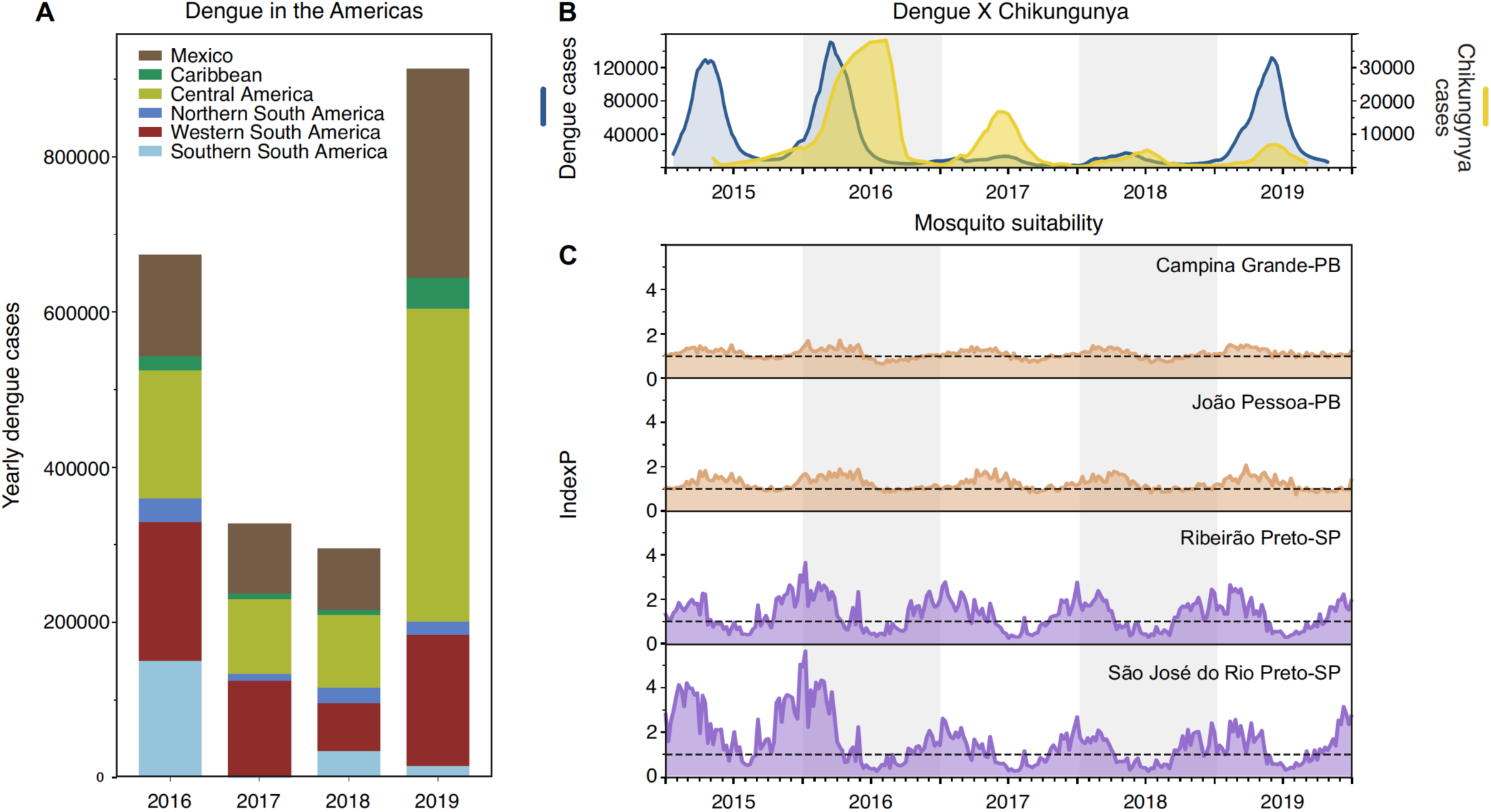
Low dengue cases in 2017-2018 not primarily due to surveillance or climate. (**A**) Dengue cases from 2016-2019 reported to the Pan-American Health Organization aggregated by region in the Americas. A sharp decrease in dengue cases was observed in 2017-2018 in distinct subcontinental regions in the Americas: Northern (from Venezuela to French Guyana), Western (from Colombia to Bolivia) and Southern South America (from Paraguay to Argentina), Caribbean, Central America, and Mexico. (**B**) In Brazil, the comparative epidemiological curves of dengue and chikungunya cases before and after major Zika outbreaks in 2016. (**C**) *Index P* (transmission potential) estimated from distinct urban areas in Paraíba (PB, northeast Brazil) and São Paulo (SP, southeast Brazil) states. No

### Virus genomics reveals the timing of the DENV outbreak lineages

When dengue incidence resurged throughout Brazil in 2019 (**Figure 1**), it was unclear if it was caused by new DENV introductions or by lineages already established in the country, which survived two years of low transmission. To investigate, we sequenced DENV from the recent outbreaks in Northeast Brazil (mainly affected by DENV-1) and Southeast Brazil (mainly affected by DENV-2)^24^, regions that typically have higher dengue cases than the rest of the country (**Figure 1C**). We sequenced 43 DENV-1 genomes collected from 2018-2019 dengue cases identified in the Northeast states of Paraíba and Alagoas. From the Southeast state of São Paulo, we sequenced three DENV-1 genomes from 2018, four DENV-2 genomes from 2010, and 19 DENV-2 genomes from 2019 (**Table S1**). The DENV samples from São Paulo were sequenced by placing serum from dengue cases on FTA filter paper cards (which thereby inactivated the virus^38^) and shipping them to Yale University (**Methods; Figure S5**), thereby highlighting how this simple method^39^ can be used to supplement virus sequencing when local capacities are limited.

To uncover the origins of the recent dengue outbreaks, we combined our 46 DENV-1 and 23 DENV-2 genomes with others that were available (see Methods), and inferred time-resolved phylogenetic trees (DENV-1 = 200 total genomes, **Figure 4**; DENV-2 = 220 total genomes, **Figure 5**; root-to-tip estimates of evolutionary rate, **Figure S6**). Our data show that the time to most recent common ancestor (tMRCA) DENV-1 viruses sequenced from Paraíba and Alagoas was mid-2012 (95% Bayesian credible interval (BCI) 2011.79-2013.33; **Figure 4**), and the tMRCA for DENV-2 viruses sequenced from São Paulo was early 2014 (95% BCI 2013.33-2014.61; **Figure 5**), which was independently confirmed^40^. As there are far more available DENV envelope protein coding sequences than whole genomes^41-43^, we repeated our analysis using 1250 DENV-1 (**Figure S7**) and 1202 DENV-2 (**Figure S8**) envelope sequences reported in previous studies^41-43^, and we found similar clustering patterns and ancestral phylogeographic origins. Overall, our data and analyses reveal a common pattern: lineages of DENV-1 and DENV-2 causing outbreaks in Southeast and Northeast Brazil were most likely introduced and descended from viruses circulating in those regions before the Zika epidemic.

### Origins and spatial dispersal of DENV-1 lineages circulating from 2012-2019

Our phylogenetic analysis revealed that DENV-1 lineages causing the 2019 outbreak in Northeast Brazil had been circulating undetected in the region since at least mid-2012 (**Figure 4**). Using near-complete genomes from DENV-1 isolates from Brazil and other countries in the Americas since the 1980s, we then explored (***1***) the genetic relatedness of DENV-1 genomes in the Northeast and Southeast, (***2***) how long the outbreak lineages have been circulating in Brazil, and (***3***) the patterns of within-region spread during the period of cryptic DENV-1 transmission.

DENV-1 was first detected in Brazil in 1981 (**Figure S7**) and, up until the 1990s, was the main serotype circulating there along with DENV-2^44^. The primary DENV-1 lineages sequenced from Brazil are classified into five clades: BR1-BR5^41,42^ (**Figure 4A, Figure S7**). Our analysis shows that the 2019 DENV-1 outbreak in Northeast Brazil, and at least one case in Southeast Brazil, was primarily caused by the BR5 lineage viruses that have been circulating in Brazil for more than a decade (tMRCA = 2005-2007; **Figure 4A**). Thus, the introduction of the BR5 lineage into Northeast Brazil around mid-2012 was likely from a domestic source. In addition, one of our sequenced DENV-1 genomes from Alagoas state (Northeast Brazil) clustered with the BR4 clade, and three of our sequenced DENV-1 genomes from São Paulo state (Southeast Brazil) clustered with the BR3 clade, indicating the continued co-circulation of multiple DENV-1 lineages in Brazil (**Figure 4A, Figure S7**).

To reconstruct the regional spread dynamics of DENV-1 lineage BR5 since its introduction into Northeast Brazil, we performed continuous phylogeographic analysis to map viral spatiotemporal spread (Figure 4B). We found that DENV-1 circulated undetected in the João Pessoa metropolitan area for at least one year through 2014 (region 1, in **Figure 4B**), before moving to the metropolitan region of Campina Grande, another important urban area in Paraíba (region 2, in **Figure 4B**). In this region, DENV-1 established local transmission chains in surrounding municipalities from 2015 to 2016 (**Figure 4B**), eventually also spreading back to Southeast Brazil (São Paulo state), where it was detected in 2019 (**Figure 4A**, in purple).

To estimate the dispersal velocity of cryptic spread, we used spatiotemporal information extracted from thousands of posterior trees inferred to investigate DENV-1 local spread (**Figure 4C-D**). We found that likely in early 2016, when Zika cases were peaking in the Northeast (**Figure 1C, Figure S1**), DENV-1 started dispersing towards the western most part of Paraíba state (region 2 to 3, **Figure 4B-C**) and reached its highest dispersion rate (69.1 Km/year in mid-2016; **Figure 4D**). It was followed by a large drop in dispersal velocity, when mainly local, short range transmissions were predominant in the region after late 2017 (**Figure 4D**). Thus, our analysis suggests that the spread of DENV-1 in the region declined alongside the Zika cases in 2017, and the outbreak in 2019 was likely caused by locally established viruses.

**Figure 4.**
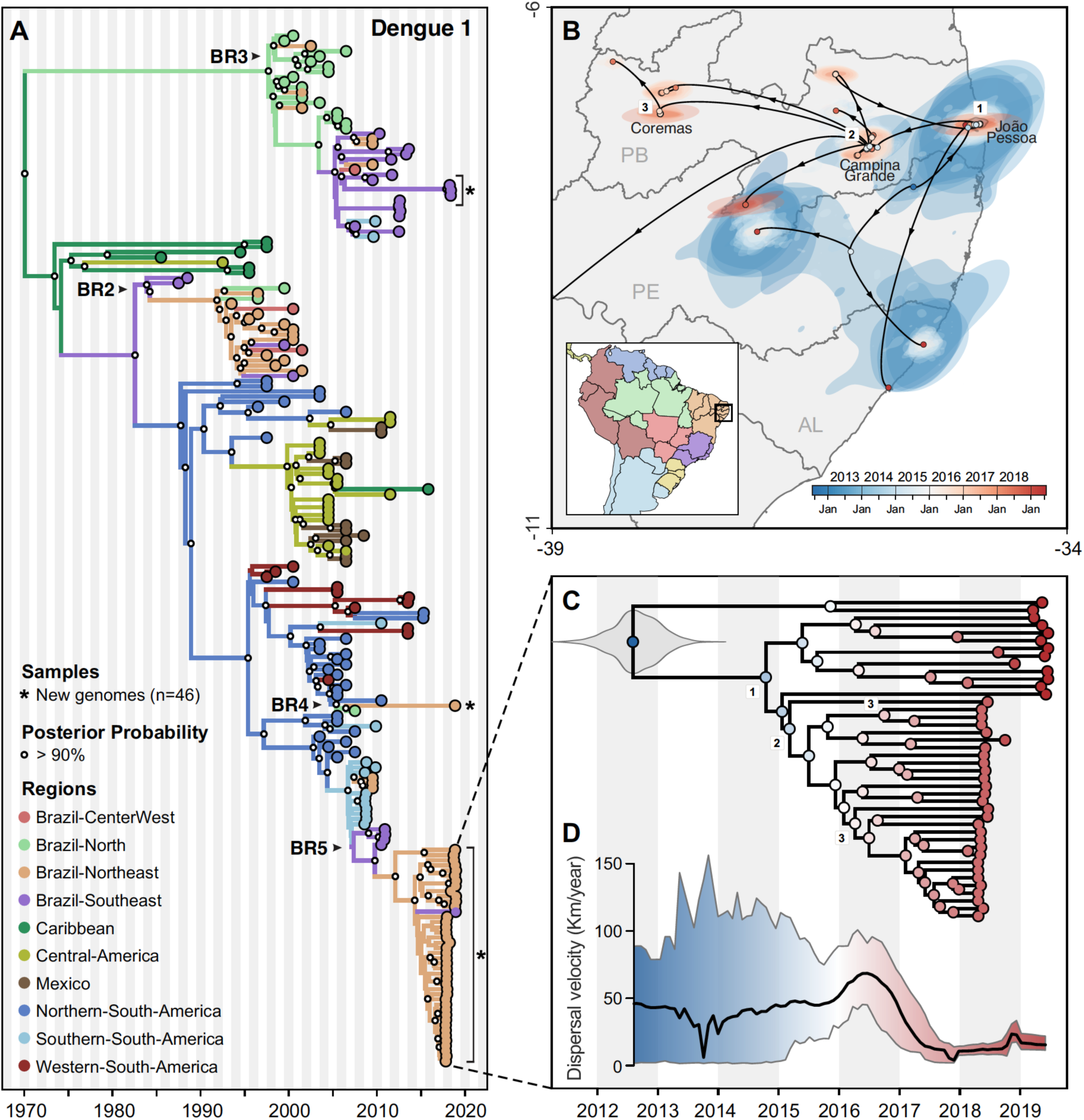
Regional emergence and cryptic transmission of DENV-1 causing recent outbreaks in Northeast Brazil. (**A**) Time-resolved phylogeny of DENV-1 circulating in the Americas since 1986 (n=200). Branch colors represent reconstructed ancestral locations using discrete phylogeography. DENV-1 genomes sequenced in this study (n=46) are highlighted with asterisks (*). (**B**) Continuous phylogeography showing the local spread of DENV-1 in the Northeast Brazil states of Paraíba (PB), Alagoas (AL), and likely Pernambuco (PE). Areas numbered as 1 (João Pessoa), 2 (Campina Grande), and 3 (Coremas) correspond to the main location where DENV-1 circulated. Shaded areas represent uncertainties, expressed as the 80% highest posterior density (HPD) of the possible locations of origin of viral ancestors. (**C**) The main DENV-1 outbreak clade plotted as movement vectors in (**B**). The violin plot shows the posterior density interval for the tMRCA. Numbers refer to areas shown in (**B**). (**D**) Weighted lineage dispersal velocity through time, reaching its peak around June 2016, with mean velocity of 69.1 km/year (confidence interval, 45.07 - 90.91 Km/year). To better depict the dynamics of spread in Northeast Brazil, the single vector leading to São Paulo was not considered in the calculations of dispersal velocity.

### Origins and spatial dispersal of DENV-2 lineages circulating from 2013-2019

Similar to DENV-1 during the outbreak in Northeast Brazil (**Figure 4**), we found that the 2019 dengue outbreak in the Southeast state of São Paulo was caused by a DENV-2 lineage established in the region since at least 2014 (**Figure 5**). To investigate the spatial dynamics of DENV-2 in Southeast Brazil, we performed similar phylogeographic analyses as described for DENV-1.

Combining our 23 DENV-2 genomes primarily from Ribeirão Preto (region 1 in **Figure 5B**) with the 20 genomes recently sequenced by de Jesus et al.^40^ from Araraquara (region 2) and Sao Jose do Rio Preto (region 3) municipalities in São Paulo state, we found the 2019 outbreak was primarily caused by a different DENV-2 lineage (BR4) than what was detected during the 2010 outbreak in the same region (BR3; **Figure 5A; Figure S8**). However, there is evidence that the DENV-2 BR3 clade, which emerged in Brazil around 2004, is still circulating in Southeast Brazil (**Figure 5A**)^40^. The closest sequenced DENV-2 ancestors to the new BR4 lineage are from the Caribbean, which was introduced into Brazil no later than 2014 (95% BCI, 2013.33 - 2014.61, **Figure 5C**).

We reconstructed the regional dispersal history of DENV-2 lineage BR4 to uncover how the virus spread in Southeast Brazil during mostly cryptic transmission (**Figure 5B-D**). Following the same approach used to investigate DENV-1 diffusion through time, we found that DENV-2 was most likely introduced in São Paulo through the municipality of Ribeirão Preto (region 1) around 2014, where it circulated undetected for 1-2 years (**Figure 5B-C**). From 2015-2017 the virus lineage reached its period of fastest dispersal (66.2 km/year) as it spread towards Araraquara and Sao Jose do Rio Preto (regions 2-3; **Figure 5C-D**). Once established in these new areas, DENV-2 circulated locally via short range, sustained transmission chains, as revealed by its low dispersal velocity through 2018-2019 (**Figure 5D**). Similar to DENV-1 lineage BR5 in Northeast Brazil, the patterns of DENV-2 lineage BR4 dispersal in Southeast Brazil followed the fall of Zika cases in Brazil (**Figure 1**), with 2019 dengue outbreaks being caused by locally established viruses rather than new introductions.

**Figure 5.**
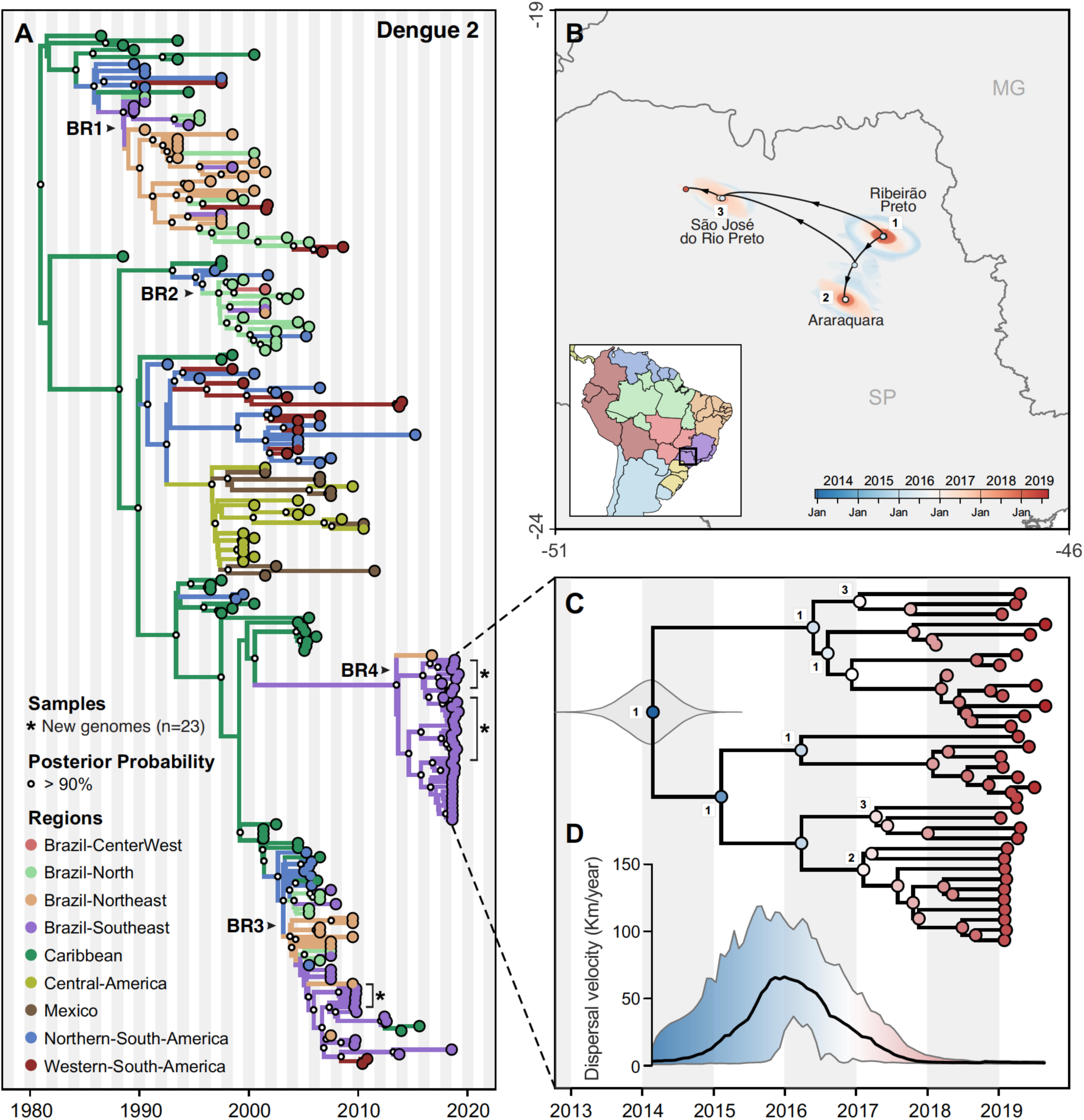
Regional emergence and cryptic transmission of dengue 2 viruses causing recent outbreaks in Southeast Brazil. (**A**) Time-resolved phylogeny of DENV-2 circulating in the Americas since 1987 (n=220). Branch colors represent reconstructed ancestral locations using discrete phylogeography. DENV-2 genomes sequenced in this study (n=23) are highlighted with asterisks (*). (**B**) Continuous phylogeography showing the local spread of DENV-2 in Southeast Brazil, state of Sao Paulo (SP). Areas numbered as 1 (Ribeirao Preto), 2 (Araraquara), and 3 (Sao Jose do Rio Preto) correspond to the main location where DENV-2 circulated. Shaded areas represent uncertainties, expressed as the 80% highest posterior density (HPD) of the possible locations of origin of viral ancestors. (**C**) Main DENV-2 outbreak clade in (**A**), plotted as movement vectors in (**B**). The violin plot shows the posterior density interval for the tMRCA. Numbers refer to areas shown in (**B**). (**D**) Weighted lineage dispersal velocity through time, reaching its peak around January 2016, with mean velocity of 66.2 km/year (confidence interval, 31.3 - 103.5 km/year).

## Discussion

### Resurgence of dengue following a period of low transmission

Following the major Zika epidemic in 2016, dengue incidence remained low throughout the Americas for two years (2017-2018), resurging in Brazil and elsewhere in 2019 (**Figure 1**). The reasons for the drastic declines in dengue cases have been debated^78^, but evidence to explain such phenomena are scarce. In this study we combined genomic, ecological and epidemiological data to explain the dynamics of dengue resurgence. We found that DENV transmission likely decreased in 2017-2018 due to low population susceptibility (**Figure 2**), rather than environmental conditions or surveillance gaps (**Figure 3**). Using viral genomes collected in 2010, 2018, and 2019, we uncovered the origins of recent dengue outbreaks in Brazil, which were caused by endemic DENV lineages circulating in Brazil for 5-10 years (**Figures 4 and 5**). This finding revealed that DENV can endure periods of low transmission, and resurge when conditions are suitable. Our results suggest that DENV cryptically circulated in 2017-2018 before resurging a year later, likely as a result of waning cross-protection conferred by prior DENV and/or ZIKV population immunity that may have played out in many countries across the Americas.

### Low population susceptibility contributed to low dengue incidence in 2017-2018

Our annual estimates of DENV FOI in the five regions of Brazil suggest that the number of susceptible individuals from 2017-2018 dropped to some of the lowest levels since 2002 (**Figure 2**). We found similar drops in susceptible individuals during other periods of low reported dengue incidence, such as 2004-2005. A major caveat of this analysis, however, is that we estimated FOI using all dengue case counts, regardless of serotype. Thus our analysis does not capture the individual transmission dynamics of the four DENV serotypes, which are known to co-circulate in Brazil at least since 20 1 0^45^. Still, transmission of all DENV serotypes was low during 2017-2018, as suggested by the low reported overall incidence.

While the decrease in dengue incidence in 2017-2018 may have resulted from population immunity building during years of high incidence (**Figure 1**), exposure to other flaviviruses could have also played a role^14^. Recent studies demonstrated that immunity from previous ZIKV infections can neutralize subsequent DENV infections by all serotypes^26^, without exacerbating the severity of secondary heterologous infections by DENV^46^. This phenomenon could have led to higher numbers of asymptomatic or mild dengue cases^47^, resulting in low reporting of cases not only in Brazil, but also in most countries in the Americas, which experienced similar patterns of presumed low dengue cases after Zika outbreaks. Although the cross-protection of ZIKV-induced immunity on DENV infections may likely wane, its modulatory effects on DENV pathogenesis could not only explain the decline of dengue in 2017-2018, but also its resurgence in the following years, all over the Americas^48^.

### Limited contribution of climate and mosquito control on dengue patterns

An important finding of our study was that environmental conditions were likely not the primary cause of the fluctuations in dengue incidence in Brazil. DENV transmission is highly dependent on climate factors, such as humidity and temperature^37^. By using climate-dependent and climate-independent data, we estimated mosquito suitability in four major urban areas in the states of Paraíba (Northeast) and São Paulo (Southeast region; **Figure 3C**) and found that conditions remained suitable for mosquito-borne virus transmission. Thus, we find it unlikely that climatic had a notable effect on the observed dengue changes during this study period.

On the other hand, human intervention on mosquito populations following the Zika epidemic could have impacted DENV transmission^8^. After WHO declared Zika a Public Health Emergency of International Concern (PHEIC) in February 2016, Brazil’s Ministry of Health allocated extra funds to implement measures of mosquito control, which included campaigns to raise awareness and provide information about self-protection against mosquito-borne viral infections^49^, and to eliminate mosquito breeding sites in vulnerable communities^50^. In this way, Brazil’s preparedness after the Zika epidemic in 2016 may account for low transmission of mosquito-borne viruses, including DENV, in the following years. This could extend to other possible outcomes of the Zika epidemic, such as changes to surveillance programs (e.g. decrease in funding and resources) or changes in human behaviour (e.g. avoidance of mosquitoes). Unfortunately, data about the mosquito control programs, surveillance strategies, and human behavior from our study areas were not available and could not be incorporated in our analyses. However, given the synchrony of dengue throughout the Americas during 2016-2020 (**Figure 3A**), the changes in incidence are more likely due to biological factors than coordinated human activities.

### Endemic dengue viruses lineages responsible for recent outbreaks

Despite the low dengue incidence in 2017-2018, we found that DENV lineages already circulating in Brazil for 5-10 years re-emerged to cause outbreaks in the Southeast and Northeast regions in 2019. This demonstrates that (!) major dengue outbreaks may not be caused by recent virus introductions and (2) DENV can survive periods of high population immunity through low-level cryptic transmission. The 2019 outbreaks caused by DENV-1 in Northeast and Southeast Brazil included at least three endemic lineages, named sequentially based on their date of introduction in Brazil^41,42^: BR3, BR4, and mainly BR5 (**Figure 4**). The 2019 outbreaks caused by DENV-2 in Southeast Brazil (São Paulo state) included the lineages BR3 and mainly BR4 (**Figure 5**), as also reported in a recent study^40^. Our continuous phylogeographic analyses for DENV-1 and DENV-2 indicate that following introduction, lineages go through periods of regional dispersal before becoming established in an urban area leading to a local outbreak.

Although our DENV genome sampling in Northeast Brazil was spatially heterogeneous, our sampling in Southeast Brazil was highly focused on two major urban areas. This may have introduced biases in our patterns of DENV spread depicted in our continuous phylogenetic analyses. Future studies investigating the genomic epidemiology of DENV, or any pathogen, should ideally consider the inclusion of genomes sampled in times and locations in such a way to match the incidence of disease to ensure a better reconstruction of the geographic patterns of spread.

### Future directions and implications

Our study challenges the paradigm that dengue outbreaks are caused by recently introduced new lineages, but rather they may be driven by established lineages circulating at low levels until the conditions are conducive for outbreaks. How DENV lineages can persist undetected through long periods of low transmission is uncertain, and should be further investigated. Furthermore, the outcomes of the present study do not directly identify what caused the fall and rise of dengue, observed in several countries^15,16,17,18^, but our data support a hypothesis pertaining to the role of the 2016 Zika epidemic and prior dengue outbreaks in this phenomenon. Further research and cohort studies are required to investigate whether previous exposure to ZIKV provides transient protection against DENV infection or severe disease outcomes, and if it has a similar effect on all DENV serotypes. As ZIKV and DENV also co-circulate in other parts of the world, understanding these immune interactions may help us to better forecast outbreaks. Furthermore, data about governmental mosquito control programs and public health campaigns should be made openly available to better evaluate the efficacy of these activities on controlling outbreaks. Finally, we advise for sustained genomic surveillance of DENV to detect cryptic lineages in an effort to plan for dengue resurgence. Collectively, our study revealed new insights into endemic DENV transmission and spread, which provides a basis for developing targeted control strategies.

## Methods

### Ethics

Human sampling from Paraiba and Alagoa states used in this study was approved for the Research Ethic Committee (REC) from the Aggeu Magalhaes Institute (IAM) under the CAAE number 10117119.6.0000.5190. Human sampling from São Paulo state used in this study was approved by REC from Hospital das Clinicas de Ribeirao Preto (HCRP) process number 12603/2006 and also by DAEP/CSE-FMRP-USP project number 19/2007. Dengue virus sequencing and analysis of a de-identified and limited dataset at the Yale School of Public Health was determined to be exempt from human research determined by a limited Yale Institutional Review Board (IRB) review (IRB protocol ID: 2000025320).

### Epidemiological data

Dengue, Zika and chikungunya case data per epidemiological week in Brazil were obtained from the Ministry of Health of Brazil, from its SINAN database ^51^ and its epidemiological bulletins ^52^, and shared by the Infectious Disease Dynamics Group at University of Florida (UF-IDD). Dengue case data per year in other countries in the Americas were obtained from PAHO ^4^. Links to all datasets can be found in Data Availability.

### Estimating force of infection

We estimated the force of infection, λ*_y,r,_* of DENV on an annual basis for each year, *y*, between 2002 and 2019 for each region, *r*. These estimates were informed by the number of cases of dengue fever, *F_y,r,a_*, and severe dengue, *S_y,r,a_*, for each year, region, and age, *a*. We calculated the likelihood of a given series of annual forces of infection in region *r*, 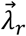, as

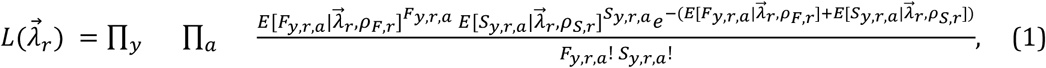

where *ϱ_F,r_* and *ϱ_s,r_* are region-specific probabilities of reporting a case of dengue fever or severe dengue, respectively, and 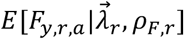 and 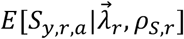 are the expected numbers of cases of dengue fever and severe dengue predicted by the model. This assumes that observed cases of each type in each year, region, and age are independent Poisson random variables.

We calculated the expected number of cases of dengue fever as

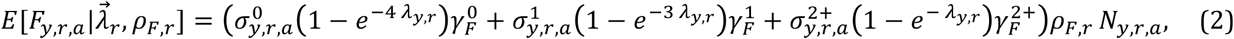

where 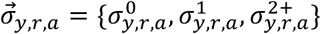 are probabilities of having experienced 0, 1, or 2+ previous DENV infections, 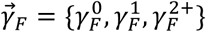 are probabilities of experiencing dengue fever conditional on infection given 0, 1, or 2+ previous DENV infections, and *N_y,r,a_* is the population of individuals of age *a* in region *r* in year *y*. The calculation for 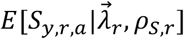 is similar.

Probabilities of having experienced a given number of previous DENV infections depend on the annual force of infection experienced in each year of a person’s life. In the first two years of life, 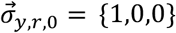 and 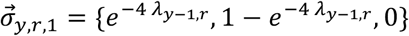 because we assume that, at most, one DENV infection is possible in each year of life. For all other ages,

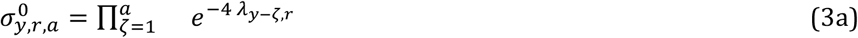

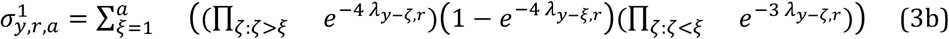

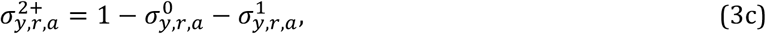

where *ζ* is an index for a year in which an individual was not infected and *ξ* is an index for a year in which an individual was infected. For 2001 and earlier, we treated force of infection as the mean of 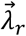. To avoid keeping track of all possible pairs of years in which an individual could have experienced two infections, we combined the proportion of individuals who had experienced two or more infections in eqn. 3c. Given that most infections resulting in dengue fever or severe dengue likely result from primary or secondary infections^53^, we viewed this approximation as acceptable. Also in the interest of model tractability, we assumed that serotypes were evenly distributed within and across years and that they were identical in terms of infectiousness, virulence, and other traits, as have previous analyses of age-stratified case data ^54^.

We used maximum-likelihood inference to estimate 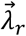 separately for each region *r* conditional on 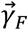, 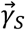, *ϱ_F,r_*, and *ϱ_s,r_*. To account for uncertainty in the latter parameters that we did not estimate, we took 100 independent, random draws of them from uniform distributions specified in **Table S2** for 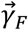 and 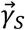. Because estimates of *ϱ_F,r_* and *ϱ_S,r_* were not readily available in the literature, we defined plausible values of *ϱ_F,r_*, and *ϱ_S,r_* as those compatible with 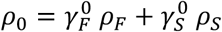, where *ρ*_0_ is defined as the probability of reporting a case resulting from a primary infection and was previously estimated to range 0.1-0.9 in Brazil ^54^. Given draws of 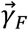 and 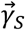 from uniform ranges defined in **Table S2** and draws of *ϱ_F,r_* and *ϱ_S,r_* compatible with 0.1 < *ρ*_0_*<* 0.9, we then obtained maximum-likelihood estimates of 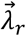. We initialized the optimization algorithm at values of

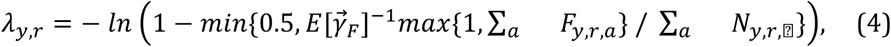

which correspond to simplified estimates based only on dengue fever cases and ignoring immunity from years preceding y.

### Mosquito transmission potential estimates

We used the Bayesian approach developed by Obolski et al. ^37^ to estimate the weekly transmission potential *(Index* P). Briefly, we obtained daily temperature and relative humidity records for four cities in Brazil from openweathermap.org. We combined these data with ecological and entomological priors documented in the literature (**Table S3**) using the R package MVSE ^37^ to estimate daily transmission potential, which we then aggregated by week.

### Clinical sample collection and virus diagnostics

#### Overview

Serum samples from dengue cases were collected from two regions in Brazil: São Paulo state in the Southeast, and; Paraíba and Alagoas states in the Northeast. Sample collection location and dates are provided in **Table S1**. All patients presented at least with fever and two or more symptoms such as nausea, vomiting, rash, myalgia, headache, retroorbital pain, petechiae or positive snare test and leukopenia.

#### Sao Paulo state

Dengue patient serum samples were collected at Hospital das Clinicas from Ribeirao Preto Medical School and at Basic Health Units (UBDS) distributed throughout the municipality. Dengue virus infection was first tested by a rapid IgM/IgG test (Dengue DUO ECO test, ECO Diagnóstica, Corinto, MG, Brazil) or NS1 ELISA (FOCUS Diagnostics, Cypress, CA, USA). Positive IgM/IgG or NS1 tests were then confirmed by RT-qPCR using the following protocol. RNA from the serum samples were extracted using KASVI (Mobius Life Science, Pinhais, PR, Brazil) according to the manufacturer’s recommendations. RT-qPCR was performed in one-step using TaqMan Fast Virus 1-Step Master Mix (Applied Biosystems), following the manufacturer’s recommendations. Primers and probes used were: FD1: 5’-GCACCAGGTGGGAAACGA-3’; RD1: 5’-TAGGAGCTTGAGGTGTTATGG-3’, DENV-1 probe: FAM-CTACAGAACATGGAACAAT-MGB and FD2: 5’-CTAAATGAAGAGCAGGACAAAAGGT-3’; RD2: 5’-ATCCATTTCCCCATCCTCTGT-3’, DENV-2 probe: FAM-TGCAAACACTCCATGGTA-MGB. The reaction was performed in the 7500 Fast Real-Time PCR System (Applied Biosystems).

#### Paraiba and Alagoas states

Dengue patient serum samples were collected by the Public Health Central Laboratory (LACEN) of each state and sent to the Arboviroses Reference Laboratory at the Aggeu Magalhães Institute - Fiocruz in Recife, Pernambuco, for viral identification. Dengue virus infections were detected by using the QIAamp Viral RNA kit (Qiagen, Hilden - Germany) to extract RNA from the serum samples followed by using a DENV-1-4 Real-Time RT-PCR Assay following the CDC’s recommendations ^55^.

### Virus genome sequencing

#### Overview

Due to limited next-generation sequencing (NGS) capacity available to the researchers in São Paulo state at the time of conducting this study, dengue clinical samples from this state were preserved on FTA filter paper cards and shipped to Yale University for metagenomic virus sequencing using an Illumina NovaSeq platform, as described below. From dengue virus samples collected in Paraíba and Alagoas states, a local Illumina MiSeq available at the Aggeu Magalhaes Institute was used for amplicon-based virus sequencing.

### Metagenomic virus sequencing from samples stored on FTA cards

#### Sao Paulo state

Frozen serum from DENV-1 and DENV-2 positive patients were thawed and 50 uL was mixed with 10-15 μL of RNA later (Ambion, USA). Around 55 - 65 μL of the mixture was spotted onto FTA Cards (Whatman™, GE Healthcare Bio-Sciences Corp, Piscataway, NJ, USA) and dried at room temperature. Each card (8,5 x 7,5 cm) was loaded with 16 different samples, respecting a minimal distance among the samples avoiding the sample cross-contamination. The cards were stored at room temperature until shipment to Yale University.

Upon receiving, FTA cards containing serum were frozen at −80 until processing. A scalpel was used to cut out and dice entire serum spots from FTA cards. The scalpel was sterilized between samples by submerging into 70% EtOH followed by submerging into 10% (v/v) bleach 3 times. To elute serum from FTA cards, diced pieces from each sample were placed in 350 μL of 1x TE, and rocked at 4°C for 16 hours. RNA was extracted from 200 μL of 1x TE elution using the Mag-Bind Viral DNA/RNA kit (Omega Bio-tek, USA) and eluted into 25 μL of water. DENV-2 samples were then screened via an RT-qPCR assay using FD2/RD2 primers with the iScript™ One-Step RT-PCR Kit With SYBR Green (Bio-Rad, USA). DENV-2 samples that produced a cycle threshold value >33 and a unique melting curve were processed for NGS, and all DENV-1 were processed.

DENV-1 and DENV-2 RNA eluted from FTA cards from São Paulo state were sequenced using an unbiased metagenomics approach on the Illumina NovaSeq at the Yale Center for Genome Analysis. To increase the overall quantity of input, we proceeded with a primer-extension pre-amplification SISPA approach to generate randomly amplified cDNA as described previously ^56,57^. Briefly, RNA was incubated with 1 μL Primer A (40pmol/μL) and 0.5ul of a 10mM dNTP mix from the SuperScript IV First-Strand Synthesis System (Thermo Fisher Scientific, USA) at 65 °C for 5 minutes. First-strand cDNA synthesis was carried out with this mixture using the SuperScript IV First-Strand Synthesis System without the addition of random hexamers according to manufacturer’s protocol. Second-strand synthesis was performed using the Sequenase Version 2.0 DNA Polymerase System (Thermo Fisher Scientific, USA) as previously described ^56^. Second-strand synthesis reactions were cleaned using 0.8:1 ratio of Mag-Bind TotalPure NGS (Omega Bio-Tek, USA) beads to volume of sample. Following two washes of beads with 70% EtOH, cDNA was eluted into nuclease water. cDNA was incubated with 2.5 μL of Primer B (40pm/μL) and amplified using Q5 HS High-Fidelity 2x MM (New England Biolabs, USA) for 15 cycles according to manufacturer’s instruction. PCR product was purified using a 0.8:1 ratio of Mag-Bind TotalPure NGS beads to volume of PCR mix. PCR products for each sample were quantified using the Qubit 1X dsDNA HS Assay Kit (Thermo Fisher, USA).

A total of 5 ng of dsDNA was used as input for library preparation using the KAPA Hyper Prep Kit (Roche, Switzerland) with NEXTflex™ Dual-Indexed DNA Barcodes. Following ligation of adaptors and barcodes, individual samples were amplified using universal Illumina primers and KAPA HiFi DNA Polymerase (Roche, Switzerland) according to manufacturer’s instructions. Libraries were purified using 1:1 ratio of Mag-Bind Total Pure NGS beads to volume of library, quantified using the Qubit 1X dsDNA HS Assay Kit, and pooled by equal concentrations. The final pool of libraries was size selected to 500-600 b.p. using the Pippin Prep (Sage Science, USA), and size distributions were confirmed using the Agilent Bioanalyzer High Sensitivity DNA Kit (Agilent Technologies, USA). Libraries were sequenced using a portion of a line on a NovaSeq at the Yale Center for Genome Analysis.

Elution and RNA extraction were attempted from a total of 96 serum samples stored on FTA cards. Of these, a total of 60 samples met the criteria for NGS (DENV1 N=7, DENV2 N=53). A total of 56 million read pairs (2×150) were generated from 65 libraries, including 5 control libraries that were introduced at the sample elution stage and library preparation stage (**Table S1**). The proportion of reads aligning to each respective DENV reference genome ranged from 0-29.9%, although the majority of samples (N=52) had less than 1% of reads passing QC aligned to the DENV reference genome. Each of the 5 control libraries had less than 15 total reads aligning to either the DENV1 or DENV2 reference genome. Of the 60 samples sequenced, a total of 25 (DENV1 N=4, DENV2 N=21) genomes meet our threshold of 70% at greater than 10X coverage to be included in our analysis (**Figure S5**).

### Amplicon-based virus sequencing

#### Paraíba and Alagoas states

Total RNA from Northeast Brazil samples were sequenced using a whole-genome tiled PCR amplicon-based approach ^58,59^. In brief, cDNA was synthesized using random hexamers (Invitrogen) and ProtoScript II Reverse Transcriptase (New England Biolabs). Then, the resulting cDNA was submitted to a multiplex PCR using primers covering the entire DENV-1 genomes designed using PrimalScheme (https://primalscheme.com/) ^58^. DENV-1 primer sequences can be found in **Table S4**. PCR reactions were performed using four separate pools to minimize primer competition, using Q5 Hot Start High-Fidelity DNA Polymerase (New England Biolabs). The resulting ~400 bp amplicons were quantified using Qubit dsDNA HS Assay Kit (Thermo Fisher Scientific Inc.), the four separate PCR reactions were pooled, and 2 ng were processed through the Nextera XT Library Prep Kit (Illumina, San Diego, CA, USA) and sequenced on the MiSeq Illumina platform from the Aggeu Magalhaes Institute - Fiocruz employing a MiSeq reagent kit V3 of 150 cycles with a paired-end strategy.

### Sequencing data processing

The goal of our bioinformatics pipeline was to generate complete or nearly complete DENV genome sequences for phylogenetic analysis using a reference-based consensus generation. Analysis was conducted on the high-performance computing clusters hosted by the Yale Center for Research Computing. Sequencing adaptors, primer sequences (for amplicon-based sequencing), and low quality reads were trimmed using Trimmomatic (version 0.39) ^60^. Following quality control, paired reads were aligned to a reference DENV1 (Accession number: JX669465.1) and DENV2 (Accession number: KP188569.1) genome using bwa (version 0.7.17) ^61^. The aligned reads were filtered and output as a.bam file using SAMtools (version 1.9) ^62^. Sorted.bam files containing only mapped, paired reads were visualized in Geneious Prime (version 2019.0.3) ^63^. Consensus sequences were generated in Geneious Prime and vcf-annotate (parameters –filter Qual=20/MinDP=100/SnpGap=20) and vcf-consensus, where a minimum of 10 reads were required to call a base at any given position in the genome and the threshold to insert ambiguous nucleotides into the consensus sequence was 75%. Regions of the genomes that did not reach the 10X depth of coverage cut-off had an “N” inserted into the consensus sequence. Final sequences that had greater than 70% of the genome with greater than 10X depth of coverage all showed to fit the expected molecular clock for viruses of their serotypes (**Figure S5**), and were used for phylogenetic analyses.

### Virus genomic data selection and alignment

To build our initial datasets, we downloaded from Genbank all complete genomes of genotypes of dengue viruses (serotype 1 and 2) commonly circulating in the Americas: DENV-1 genotype V (n=458), and DENV-2 genotype AA (n=814) complete genomes. Genomes without collection dates and location information were excluded. The sequences were aligned using MAFFT ^64^, manually curated and had their untranslated regions (UTR) trimmed. Preliminary maximum-likelihood (ML) analyses were performed using IQTree v.1.6.12 ^65^, with automatic model selection by the software. Using TempEst ^66^, we inspected these genomes to identify major molecular clock outliers, which were removed from all downstream analyses. This yielded two high quality datasets with genomes evenly sampled through time, allowing optimal clock signal (**Figure S6**): DENV-1 (n=458 genomes), and DENV-2 (n=700 genomes). Based on these samples, smaller datasets were obtained by subsampling overrepresented clades, keeping only two representatives per deme, and adding our newly sequenced genomes.

### Discrete phylogeographic analyses

To infer the evolutionary history of DENV-1 and 2, and understand the recent major outbreaks of dengue in Brazil, we performed Bayesian phylogenetic inference using BEAST v.1.10.4 ^67^. For these analyses we specified a Bayesian skygrid coalescent tree prior with 50 grid points ^68^, a GTR + T4 model of nucleotide substitution, and a strict molecular clock. Geographic data of sample origins were arranged in a geographic scheme of subnational (the five regions in Brazil) and subcontinental areas, such as Northern (from Venezuela to French Guyana), Western (from Colombia to Bolivia) and Southern South America (from Paraguay to Chile/Argentina), Caribbean, Central America and Mexico. The ancestral origins of viruses circulating in the Americas were inferred using a reversible discrete phylogeography model ^69^. Using the BEAGLE library (v3.1.0) to accelerate computation ^70^, Markov chain Monte Carlo (MCMC) sampling was performed for 100 million iterations, and convergence of parameters (ESS > 200) was assessed using Tracer v1.7 ^71^. After removing 10% burn-in, maximum clade credibility (MCC) trees were summarized using TreeAnnotator v1.10.4, visualized using FigTree v1.4.4 ^72^, and plotted using baltic ^73^.

### Continuous phylogeographic analyses

To infer the dispersal history of dengue virus lineages circulating in Northeast and Southeast Brazil in recent years, we used a continuous phylogeographic method^74^ implemented in BEAST v.1.10.4 ^67^. These analyses focused on two monophyletic outbreak clades of DENV-1 and 2 genomes sequenced in this study (**Figure 4C and Figure 5C**). We used the same evolutionary model as described above, using the inferred heights (95% HPD) from such analyses (**Figures 4A and 5A**) as normal prior distributions to set the tMRCA of the outbreak clades. Taking advantage of geographic coordinates associated with the outbreak viruses, a Cauchy relaxed random walk (RRW) diffusion model was employed to infer coordinates associated with ancestors of viruses causing the recent outbreaks (internal nodes). Markov chain Monte Carlo (MCMC) sampling was performed for 150 million iterations, using the BEAGLE library (v3.1.0) to accelerate computation ^70^, and convergence was inspected using Tracer v1.7 ^71^. As a result, we obtained 10,000 trees with ancestral coordinates annotated at each node. After discarding 10% of the sampled trees as burn-in, the data recorded in 1,000 trees were extracted and plotted in geographic space using the “seraphim” R package ^75^.

## Data Availability

The genomes generated in this study are available on NCBI (accession numbers: will be provided in the final submission; genomic data can be found in our github repository), and datasets used in our analysis can be found at https://github.com/grubaughlab/DENV-genomics/tree/master/paper1.

## Author contribution

Designed the study: AFB, TAP, RFOF, GLW, and NDG. Performed the sequencing: LCM, MJLS, JRF, RDOC, and CCK. Data Curation: AFB, JRF, RJO, MEP, EA, GCE, ATH, and DATC. Analyzed the data: AFB, JRF, RJO, MEP, GCE, GB, and TAP. Provided clinical samples and/or resources: LCM, MJLS, RDOC, FZD, MRP, LACJ, ECMM, LMRP, RFOF, BALF, and GLW. Writing - Original Draft Preparation: AFB, JRF, RJO, MEP, TAP, and NDG. Writing - Review & Editing: AFB, RFOF, TAP, GLW, and NDG. Supervision: GB, RFOF, TAP, BALF, GLW, and NDG.

## Acknowledgements

We acknowledge members of the Grubaugh Lab, S. Taylor, and P. Jack for their ongoing discussions about this manuscript. This work was funded by the Hetch Award provided by the Yale Institute for Global Health (NDG), the Interne Fondsen KU Leuven / Internal Funds KU Leuven under grant agreement C14/18/094 (GB), and the Research Foundation - Flanders (‘Fonds voor Wetenschappelijk Onderzoek - Vlaanderen’, G0E1420N; GB).

## Supplemental Information

**Table S1. DENV-1 and DENV-2 genomes sequenced in this study**. (Excel)

**Table S2. Parameter ranges for 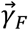 and 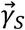 used in estimating force of infection**. (pdf)

**Table S3. Priors for Index P Estimates**. (pdf)

**Table S4. Primers for DENV-1 amplicon-based sequencing**. (Excel)

**Figure S1 Zika cases in Brazil per state, 2016-2018**. (pdf)

**Figure S2. Ranking of Force of Infection estimates**. (pdf)

**Figure S3. Probability of dengue fever (left) or severe dengue (right) conditional on DENV infection**.

**Figure S4. Proportion of populations susceptible to DENV**. (pdf)

**Figure S5. Genome coverage of DENV-1 and DENV-2 samples sequenced from FTA filter paper cards**. (pdf)

**Figure S6. Root-to-tip analyses of DENV-1 and DENV-2 genomes used in phylogenetic analyses**. (pdf)

**Figure S7. Maximum Likelihood phylogeographic reconstruction of DENV-1 evolution in the Americas using envelope sequences**. (pdf)

**Figure S8. Maximum Likelihood phylogeographic reconstruction of DENV-2 evolution in the Americas using envelope sequences**. (pdf)

**Table S2.**
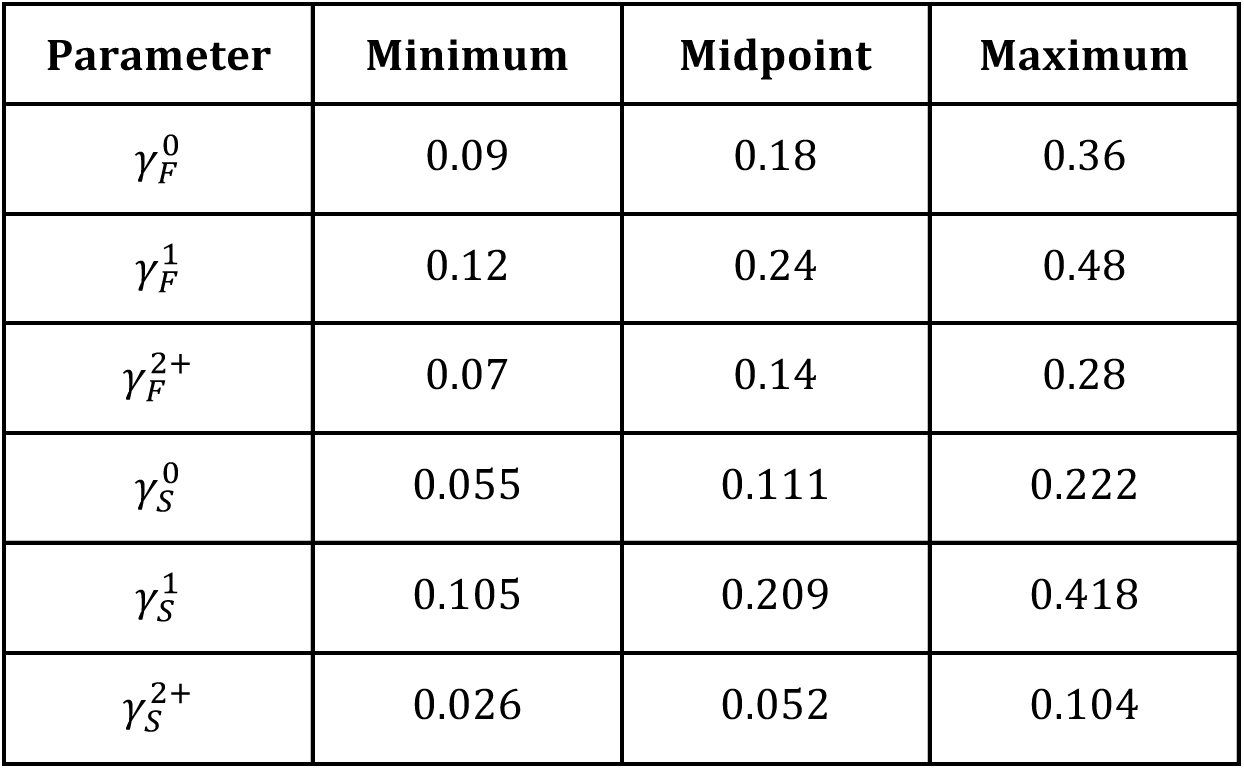
Parameter ranges for 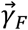 and 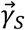 used in estimating force of infection. The midpoints of 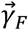 were taken from Perkins et al.^76^, and the midpoints of 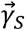 were taken from Flasche et al. ^77^. Minimum and maximum values of the range explored are 50% lower and higher, respectively, than the midpoint.

| Parameter | Minimum | Midpoint | Maximum |
| --- | --- | --- | --- |
| $\gamma_F^0$ | 0.09 | 0.18 | 0.36 |
| $\gamma_F^1$ | 0.12 | 0.24 | 0.48 |
| $\gamma_F^{2+}$ | 0.07 | 0.14 | 0.28 |
| $\gamma_S^0$ | 0.055 | 0.111 | 0.222 |
| $\gamma_S^1$ | 0.105 | 0.209 | 0.418 |
| $\gamma_S^{2+}$ | 0.026 | 0.052 | 0.104 |

**Table S3.** Priors for Index P Estimates.

| Parameter | Prior distribution | Sources |
| --- | --- | --- |
| Human Incubation Period | Gaussian (mean = 5, SD = 1) | 78-80 |
| Human Infectious Period | Gaussian (mean = 5, SD = 1) | 78-80 |
| Human Life Expectancy | Gaussian (mean = 73, SD = 2) | 81 |
| Transmission probability (human to mosquito) | Gaussian (mean = 0.5, SD = 0.01) | 37 |
| Mosquito Biting Rate | Gaussian (mean = 0.25, SD = 0.05) | 82,83 |
| Extrinsic Incubation Period | Gaussian (mean = 7, SD = 2) | 79,84,85 |
| Mosquito Life Expectancy | Gaussian (mean = 14, SD = 3) | 86,87 |

**Figure S1.**
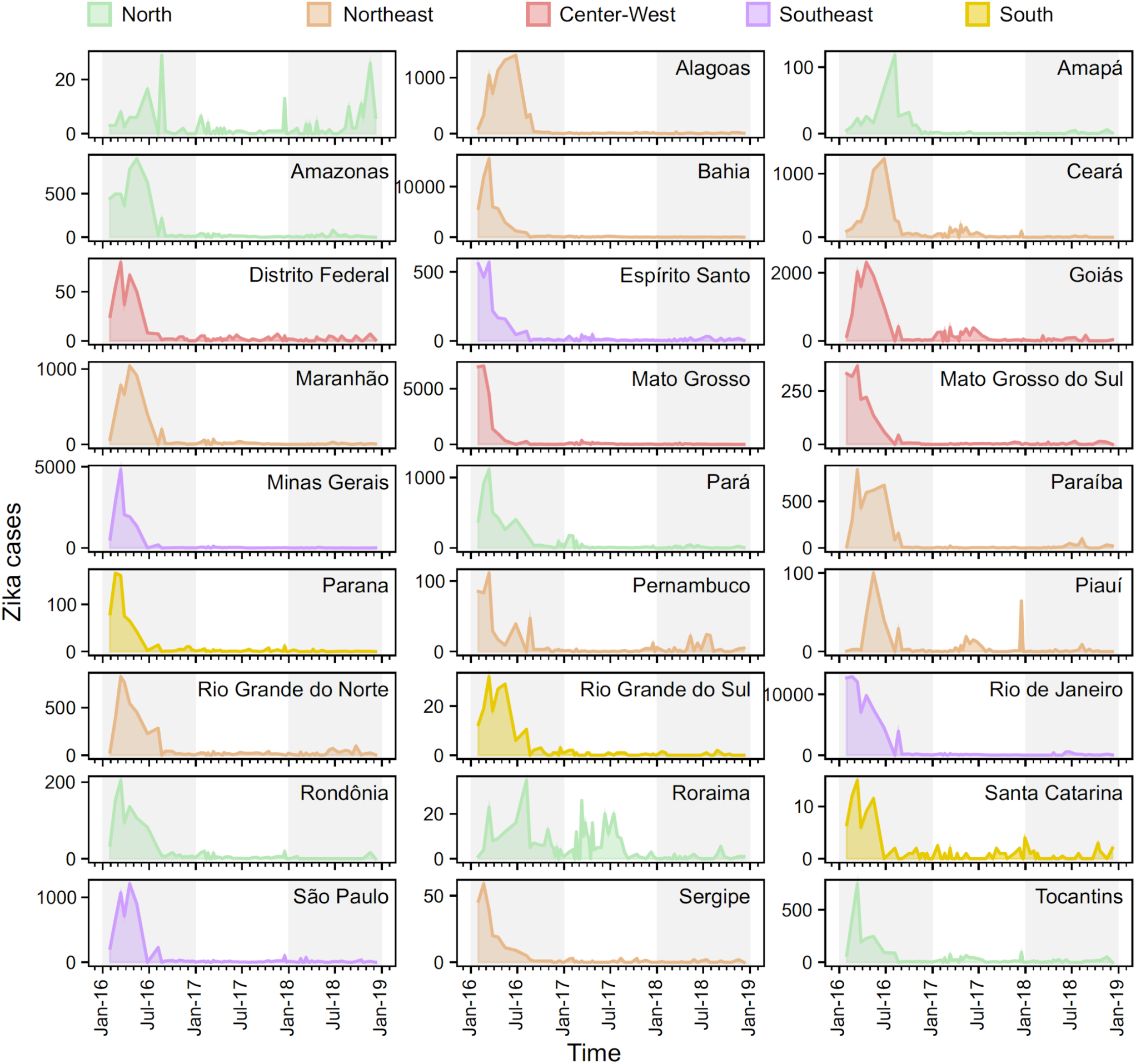
Zika cases in Brazil per state, 2016-2018. Most states experienced a peak in Zika cases between February and July 2016.

**Figure S2.**
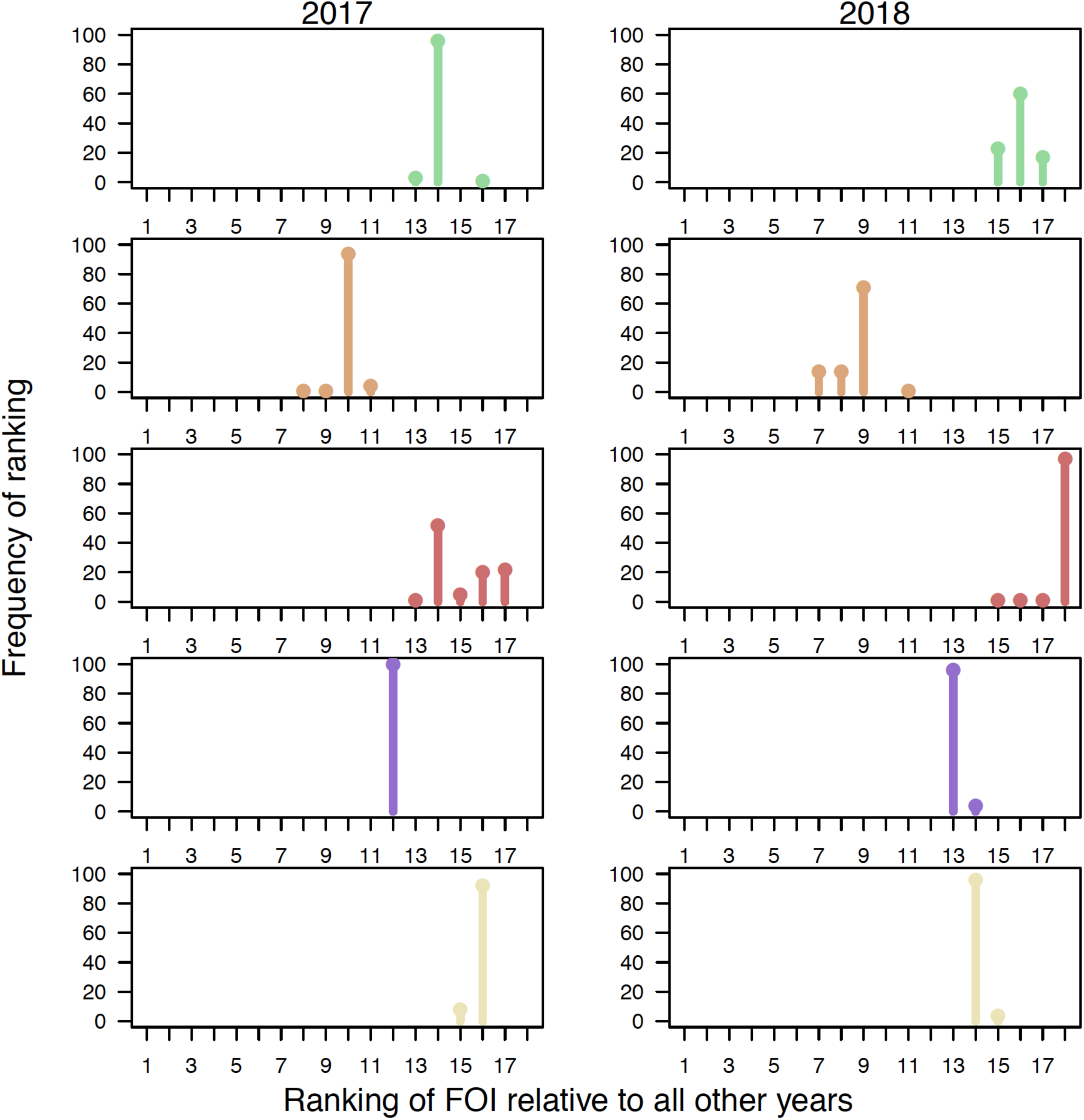
Ranking of Force of Infection estimates. Ranking 100 maximum likelihood estimates of Force of Infection (FOI) for each region in Brazil, for the two recent years of low dengue incidence (2017-2018).

**Figure S3.**
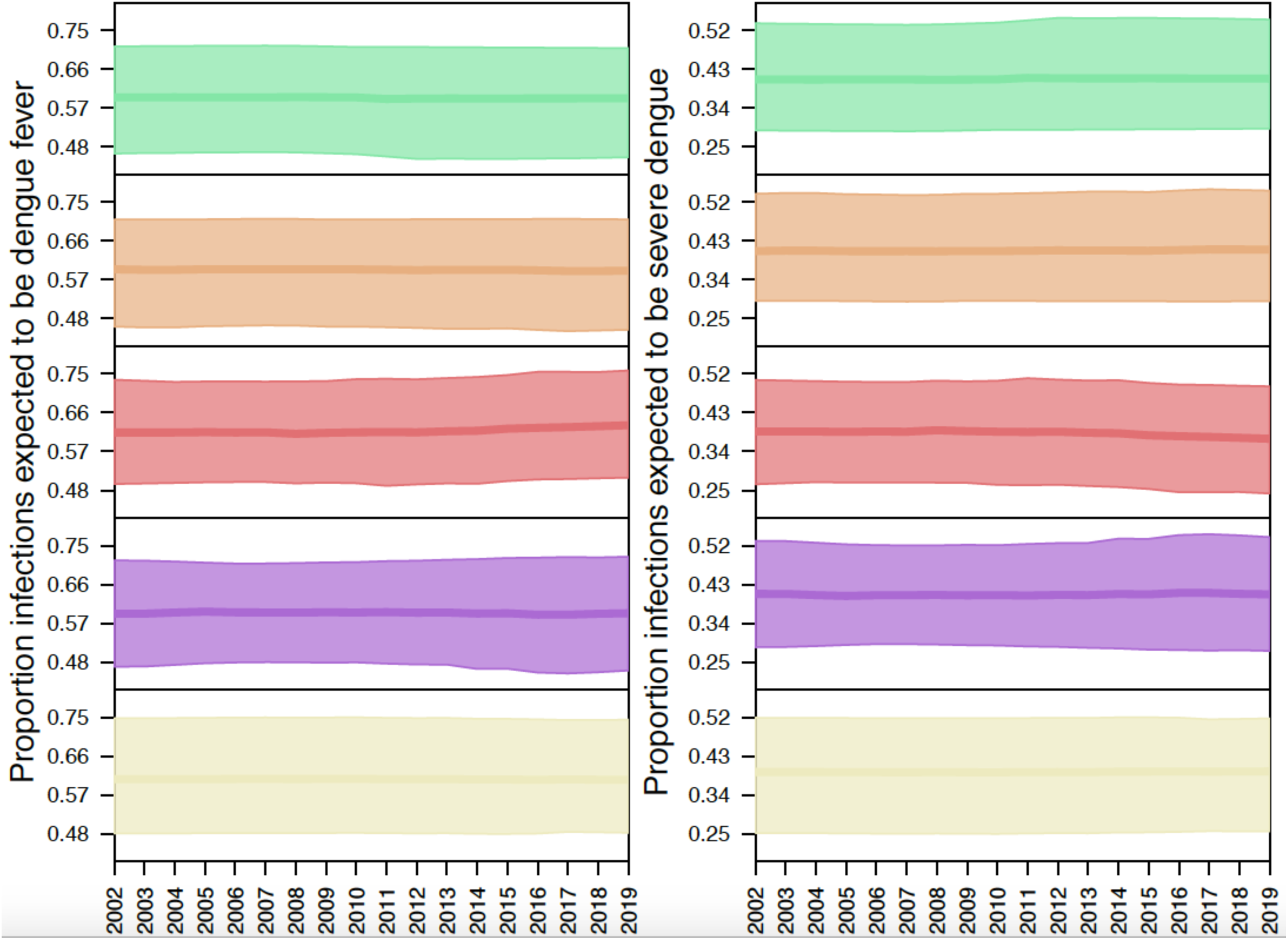
Probability of dengue fever (left) or severe dengue (right) conditional on DENV infection. Estimates are presented per geographic region in Brazil, from 2002 to 2019.

**Figure S4.**
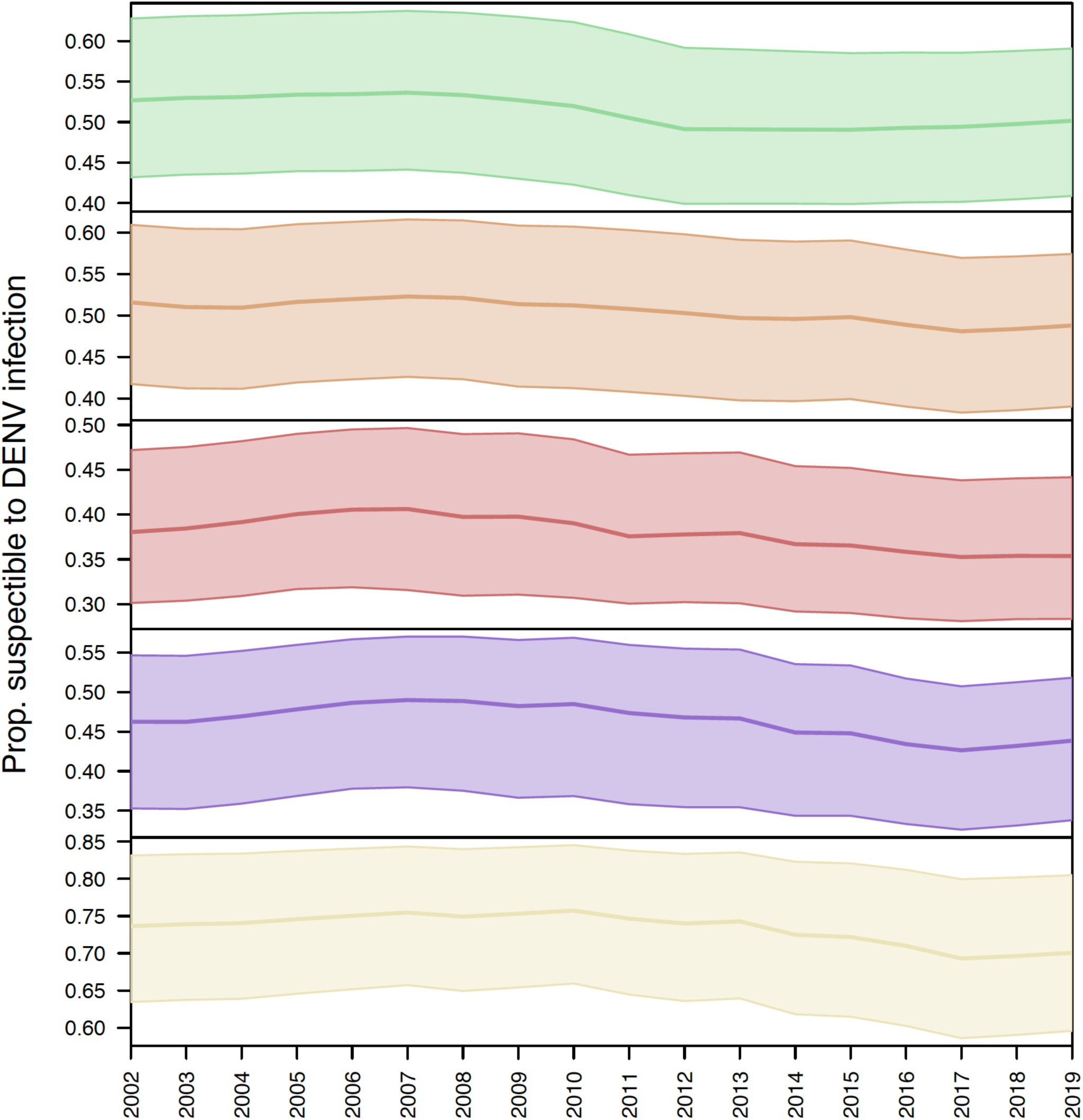
Proportion of populations susceptible to DENV. Annual proportions of susceptible population per geographic region in Brazil, from 2002 to 2019.

**Figure S5.**
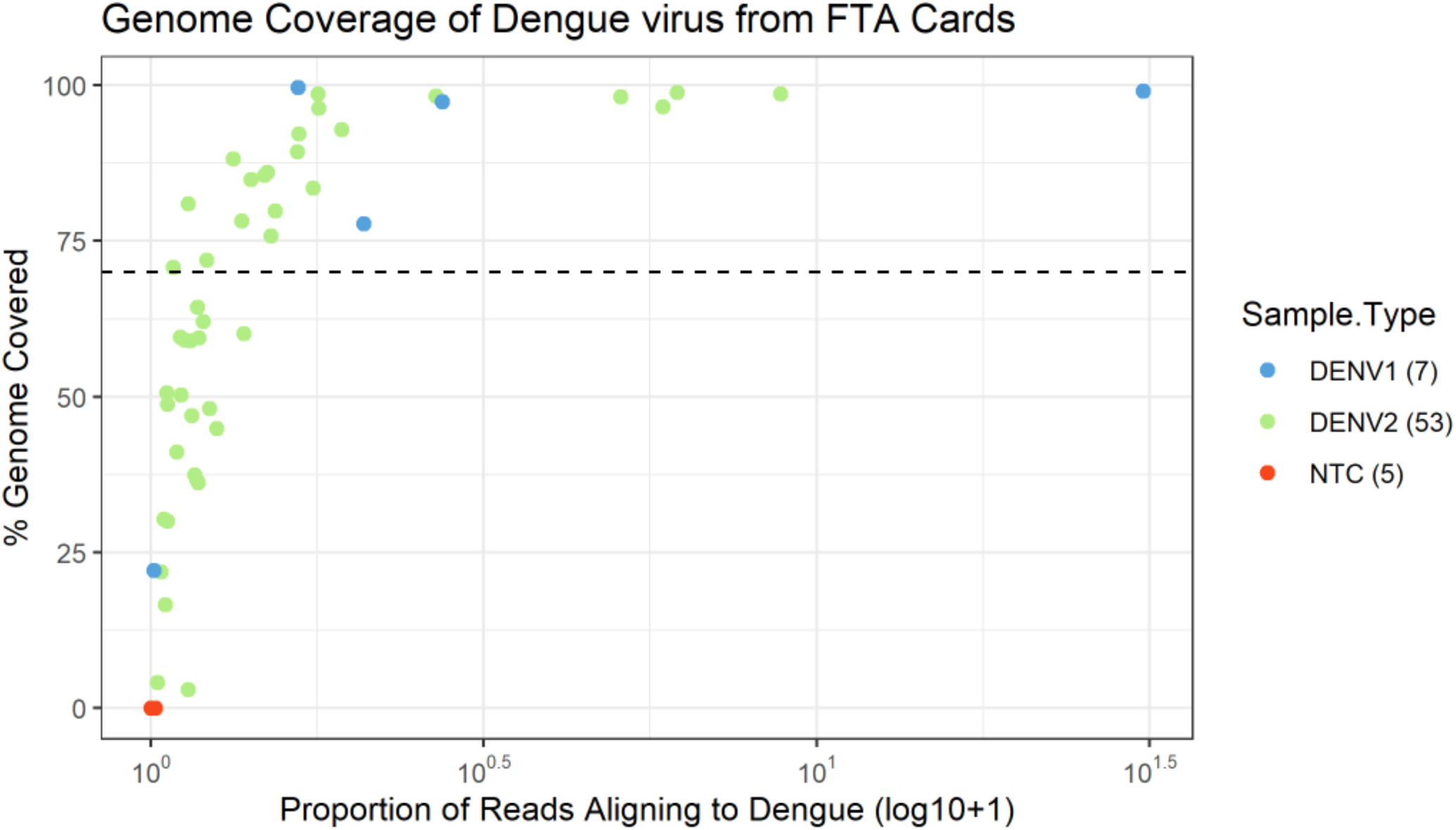
Genome coverage of DENV-1 and DENV-2 samples sequenced from FTA filter paper cards. Dotplot showing the percentage of the genome covered by >10X depth by the overall proportion of reads aligning to the DENV genome generated. Dashed line represents a 70% genome coverage threshold.

**Figure S6.**
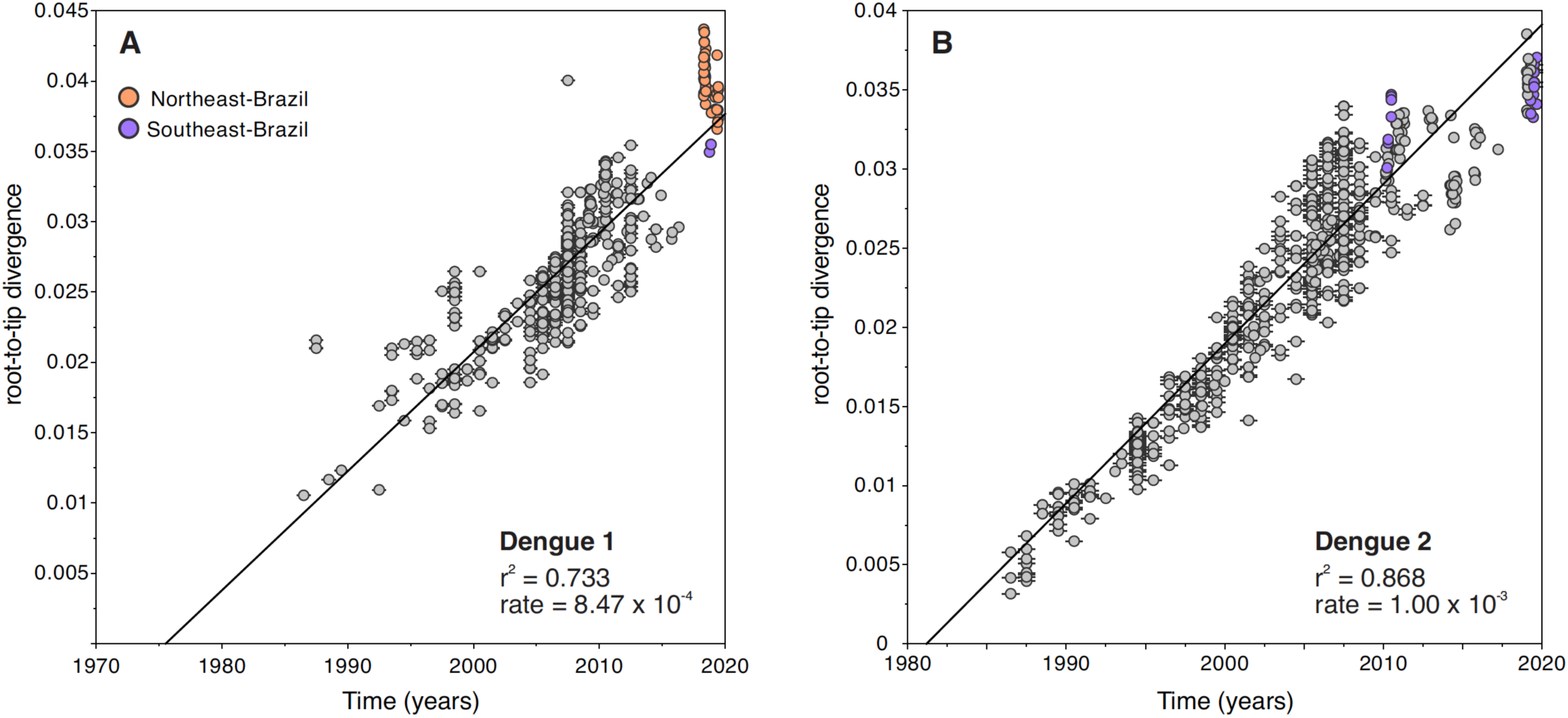
Root-to-tip analyses of DENV-1 and DENV-2 genomes used in phylogenetic analyses. These plots were obtained using TempEst, using ML phylogenies obtained from complete genomes of (A) DENV-1 (n=458) and (B) DENV-2 (n=700). They show the correlation between the collection time (years) and the genetic divergence (substitutions per site) from the root of the tree (Most Recent Common Ancestors, MRCA) to the tips (sampled genomes). The slope represents the evolutionary rate: 8.47 x 10^-4^ substitutions/site/year for DENV-1; and 1 x 10^-3^ substitutions/site/year for DENV-2. All the newly sequenced genomes generated in this study fit the expected molecular clock of viruses from their serotypes.

**Figure S7.**
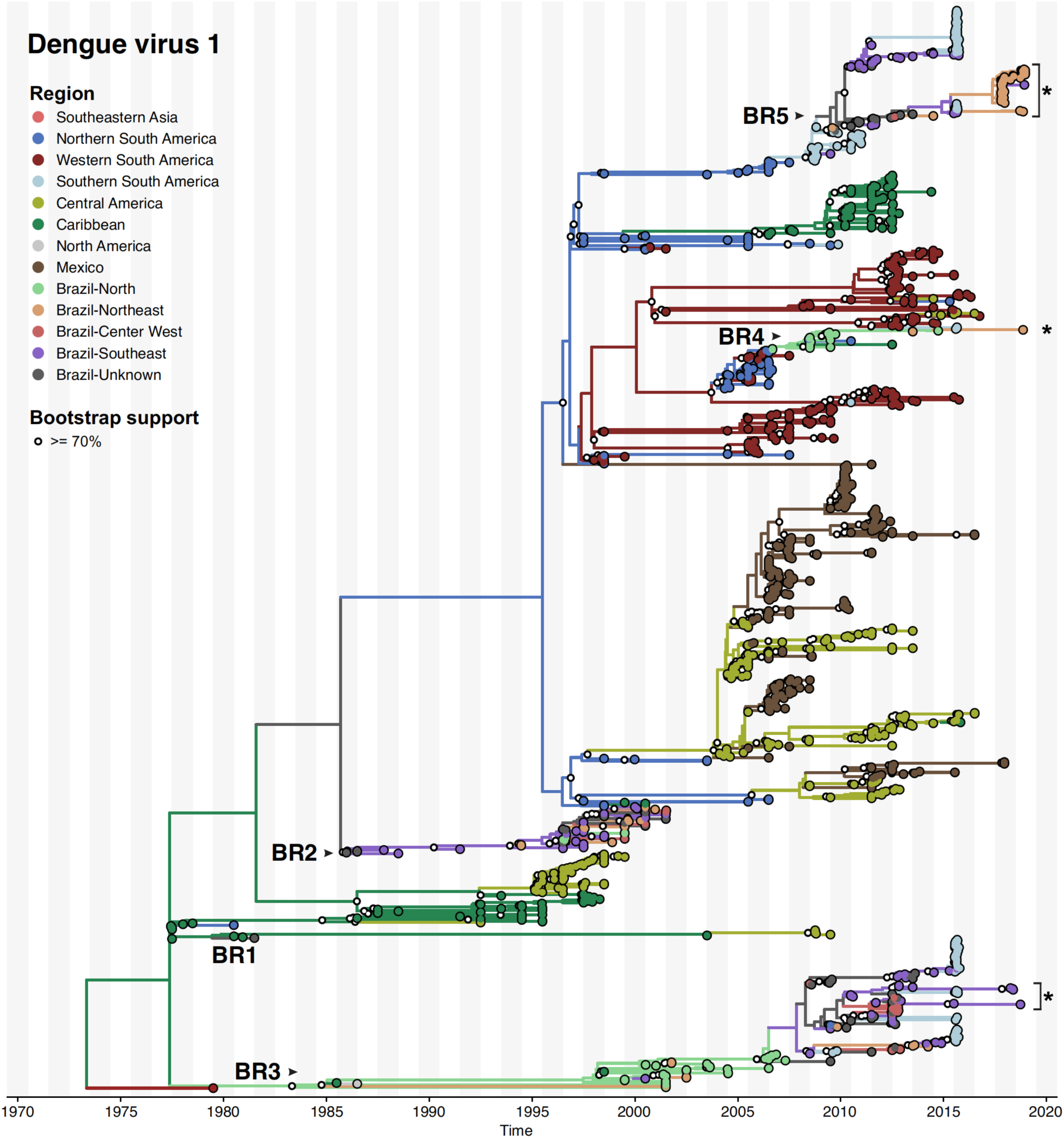
Maximum-likelihood phylogeographic reconstruction of DENV-1 evolution in the Americas using envelope sequences. A total of 1250 envelope sequences were included in this analysis (new genomes highlighted with asterisks), performed using the augur pipeline from Nextstrain. Data visualization was obtained using baltic.py. An interactive version of this phylogeographic reconstruction and metadata can be found at https://nextstrain.org/community/grubaughlab/DENV-genomics/DENV1-Brazil.

**Figure S8.**
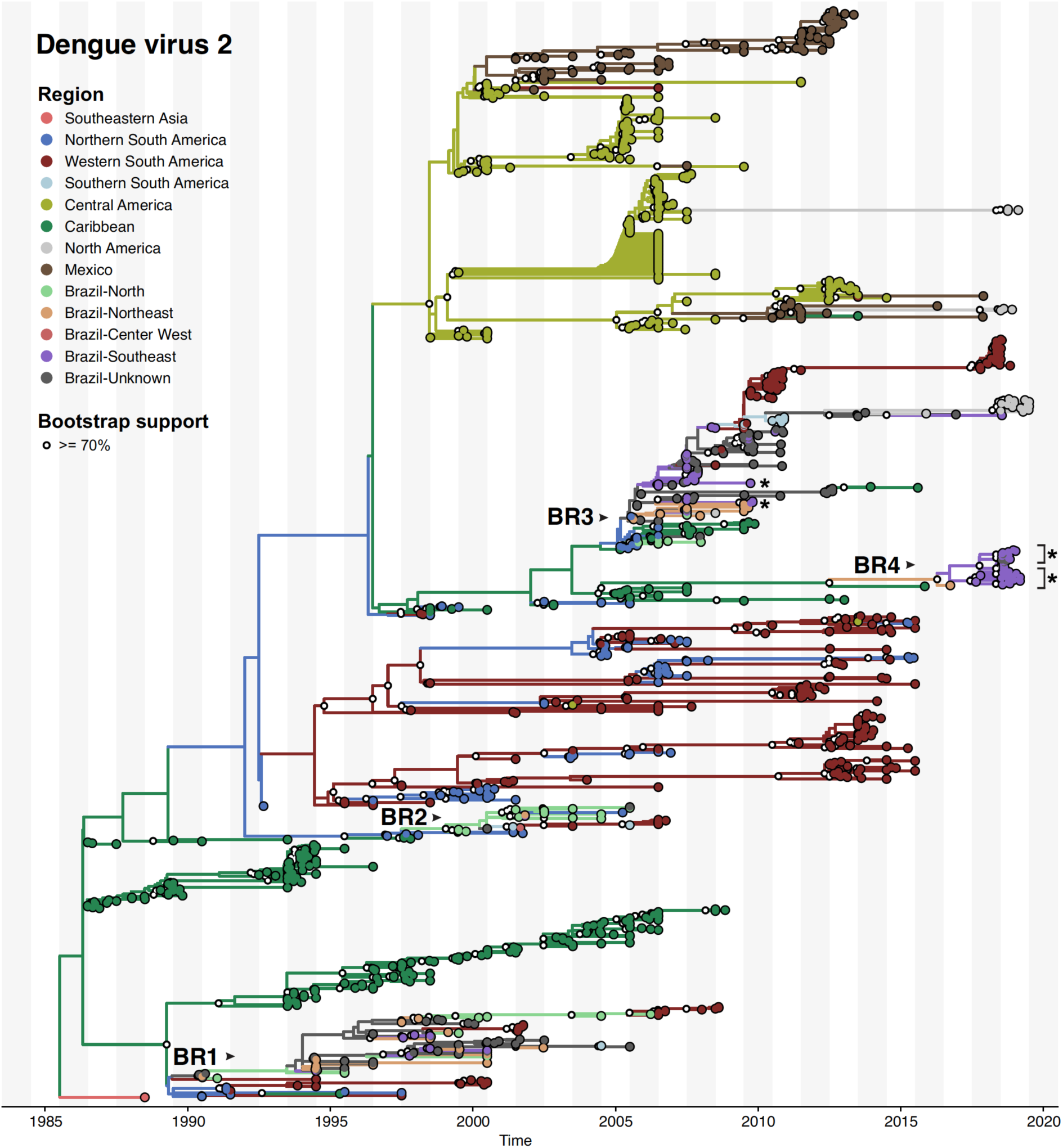
Maximum-likelihood phylogeographic reconstruction of DENV-2 evolution in the Americas using envelope sequences. A total of 1202 envelope sequences were included in this analysis (new genomes highlighted with asterisks), performed using the augur pipeline from Nextstrain. Data visualization was obtained using baltic.py. An interactive version of this phylogeographic reconstruction and metadata can be found at https://nextstrain.org/community/grubaughlab/DENV-genomics/DENV2-Brazil.

## References

1. Faria, N. R. et al. Establishment and cryptic transmission of Zika virus in Brazil and the Americas. Nature 546, 406–410 (2017).

2. Grubaugh, N. D., Faria, N. R., Andersen, K. G. & Pybus, O. G. Genomic Insights into Zika Virus Emergence and Spread. Cell 172, 1160–1162 (2018).

3. PAHO. Epidemiological Alert: Dengue. (2018).

4. PAHO. PAHO/WHO Data. Pan American Health Organization https://www.paho.org/data/index.php/en/mnu-topics/indicadores-dengue-en/dengue-nacional-en/252-dengue-pais-ano-en.html (2020).

5. Messina, J. P. et al. Mapping global environmental suitability for Zika virus. Elife 5, (2016).

6. Bhatt, S. et al. The global distribution and burden of dengue. Nature 496, 504–507 (2013).

7. Borchering, R. K. et al. Impacts of Zika emergence in Latin America on endemic dengue transmission. Nat. Commun. 10, 5730 (2019).

8. Perez, F. et al. The decline of dengue in the Americas in 2017: discussion of multiple hypotheses. Tropical Medicine & International Health vol. 24 442–453 (2019).

9. Brady, O. J. et al. Global temperature constraints on Aedes aegypti and Ae. albopictus persistence and competence for dengue virus transmission. Parasit. Vectors 7, 338 (2014).

10. Halstead, S. B. Dengue antibody-dependent enhancement: knowns and unknowns. Microbiol Spectr. 2, (2014).

11. Castanha, P. M. S. et al. Reciprocal immune enhancement of dengue and Zika virus infection in human skin. JCI insight 5, (2020).

12. Halstead, S. B. Dengue Antibody-Dependent Enhancement: Knowns and Unknowns. Microbiol Spectr 2, (2014).

13. Oliveira, R. A. et al. Previous dengue or Zika virus exposure can drive to infection enhancement or neutralisation of other flaviviruses. Mem. Inst. Oswaldo Cruz 114, e190098 (2019).

14. Ribeiro, G. S. et al. Does immunity after Zika virus infection cross-protect against dengue? Lancet Glob Health 6, e140–e141 (2018).

15. Netto, E. M. et al. High Zika Virus Seroprevalence in Salvador, Northeastern Brazil Limits the Potential for Further Outbreaks. mBio vol. 8 (2017).

16. Villarroel, P. M. S. et al. Zika virus epidemiology in Bolivia: A seroprevalence study in volunteer blood donors. PLoSNegl. Trop. Dis. 12, e0006239 (2018).

17. Langerak, T. et al. Zika Virus Seroprevalence in Urban and Rural Areas of Suriname, 2017. J. Infect. Dis. 220, 28–31 (2019).

18. Zambrana, J. V. et al. Seroprevalence, risk factor, and spatial analyses of Zika virus infection after the 2016 epidemic in Managua, Nicaragua. Proc. Natl. Acad. Sci. U. S. A. 115, 9294–9299 (2018).

19. Campos, T. D. L. et al. Revisiting Key Entry Routes of Human Epidemic Arboviruses into the Mainland Americas through Large-Scale Phylogenomics. Int. J. Genomics Proteomics 2018, 6941735 (2018).

20. Carrington, C. V. F., Foster, J. E., Pybus, O. G., Bennett, S. N. & Holmes, E. C. Invasion and maintenance of dengue virus type 2 and type 4 in the Americas. J. Virol. 79, 14680–14687 (2005).

21. Foster, J. E., Bennett, S. N., Carrington, C. V. F., Vaughan, H. & McMillan, W. O. Phylogeography and molecular evolution of dengue 2 in the Caribbean basin, 1981-2000. Virology 324, 48–59 (2004).

22. Allicock, O. M. et al. Phylogeography and population dynamics of dengue viruses in the Americas. Mol. Biol. Evol. 29, 1533–1543 (2012).

23. de Araújo, J. M. G., Bello, G., Romero, H. & Nogueira, R. M. R. Origin and evolution of dengue virus type 3 in Brazil. PLoS Negl. Trop. Dis. 6, e1784 (2012).

24. Brasil. Boletim Epidemiológico. (2019).

25. McElroy, K. L. et al. Endurance, refuge, and reemergence of dengue virus type 2, Puerto Rico, 1986-2007. Emerg. Infect. Dis. 17, 64–71 (2011).

26. Perez-Guzman, E. X. et al. Time elapsed between Zika and dengue virus infections affects antibody and T cell responses. Nat. Commun. 10, 4316 (2019).

27. PAHO/WHO Data - Dengue. https://www.paho.org/data/index.php/en/mnu-topics/indicadores-dengue-en.html.

28. Hennessey, M., Fischer, M. & Erin Staples, J. Zika Virus Spreads to New Areas — Region of the Americas, May 2015-January 2016. MMWR. Morbidity and Mortality Weekly Report vol. 65 1–4 (2016).

29. Clapham, H. E., Cummings, D. A. T. & Johansson, M. A. Immune status alters the probability of apparent illness due to dengue virus infection: Evidence from a pooled analysis across multiple cohort and cluster studies. PLoSNegl. Trop. Dis. 11, e0005926 (2017).

30. Adams, B. et al. Cross-protective immunity can account for the alternating epidemic pattern of dengue virus serotypes circulating in Bangkok. Proc. Natl. Acad. Sci. U. S. A. 103, 14234–14239 (2006).

31. Wearing, H. J. & Rohani, P. Ecological and immunological determinants of dengue epidemics. Proc. Natl. Acad. Sci. U. S. A. 103, 11802–11807 (2006).

32. Cummings, D. A. T. et al. The impact of the demographic transition on dengue in Thailand: insights from a statistical analysis and mathematical modeling. PLoS Med. 6, e1000139 (2009).

33. van Panhuis, W. G. et al. Region-wide synchrony and traveling waves of dengue across eight countries in Southeast Asia. Proc. Natl. Acad. Sci. U. S. A. 112, 13069–13074 (2015).

34. Churakov, M., Villabona-Arenas, C. J., Kraemer, M. U. G., Salje, H. & Cauchemez, S. Spatio-temporal dynamics of dengue in Brazil: Seasonal travelling waves and determinants of regional synchrony. PLoS Negl. Trop. Dis. 13, e0007012 (2019).

35. Cazelles, B., Chavez, M., McMichael, A. J. & Hales, S. Nonstationary influence of El Niño on the synchronous dengue epidemics in Thailand. PLoS Med. 2, e106 (2005).

36. Hoang Quoc, C. et al. Synchrony of Dengue Incidence in Ho Chi Minh City and Bangkok. PLoS Negl. Trop. Dis. 10, e0005188 (2016).

37. Obolski, U. et al. MVSE: An R-package that estimates a climate-driven mosquito-borne viral suitability index. Methods Ecol. Evol. 10, 1357–1370 (2019).

38. Smit, P. W., Elliott, I., Peeling, R. W., Mabey, D. & Newton, P. N. An overview of the clinical use of filter paper in the diagnosis of tropical diseases. Am. J. Trop. Med. Hyg. 90, 195–210 (2014).

39. Grubaugh, N. D. et al. Xenosurveillance: a novel mosquito-based approach for examining the human-pathogen landscape. PLoS Negl. Trop. Dis. 9, e0003628 (2015).

40. de Jesus, J. G. et al. Genomic detection of a virus lineage replacement event of dengue virus serotype 2 in Brazil, 2019. Mem. Inst. Oswaldo Cruz 115, e190423 (2020).

41. Dutra, K. R. et al. Molecular surveillance of dengue in Minas Gerais provides insights on dengue virus 1 and 4 circulation in Brazil. J. Med. Virol. 89, 966–973 (2017).

42. de Bruycker-Nogueira, F., Mir, D., Dos Santos, F. B. & Bello, G. Evolutionary history and spatiotemporal dynamics of DENV-1 genotype V in the Americas. Infect. Genet. Evol. 45, 454–460 (2016).

43. Drumond, B. P. et al. Circulation of different lineages of Dengue virus 2, genotype American/Asian in Brazil: dynamics and molecular and phylogenetic characterization. PLoS One 8, e59422 (2013).

44. Villabona-Arenas, C. J. & Zanotto, P. M. de A. Worldwide spread of Dengue virus type 1. PLoS One 8, e62649 (2013).

45. Salles, T. S. et al. History, epidemiology and diagnostics of dengue in the American and Brazilian contexts: a review. Parasit. Vectors 11, 264 (2018).

46. Breitbach, M. E. et al. Primary infection with dengue or Zika virus does not affect the severity of heterologous secondary infection in macaques. PLoS Pathog. 15, e1007766 (2019).

47. do Nascimento, I. D. S. et al. Retrospective cross-sectional observational study on the epidemiological profile of dengue cases in Pernambuco state, Brazil, between 2015 and 2017. BMC Public Health 20, 923 (2020).

48. Ribeiro, G. S. et al. Influence of herd immunity in the cyclical nature of arboviruses. Curr. Opin. Virol. 40, 1–10 (2020).

49. Brasil. Protocolo de atenμao a saude e resposta a ocorrencia de microcefalia relacionada a infecμao pelo virus zika. Ministerio da Saude, Secretaria de Atenμao a Saude http://www.saude.gov.br/images/pdf/protocolo-sas-2.pdf (2016).

50. Gómez, E. J., Perez, F. A. & Ventura, D. What explains the lacklustre response to Zika in Brazil? Exploring institutional, economic and health system context. BMJ Glob Health 3, e000862 (2018).

51. Brasil. DATASUS. DATASUS http://www2.datasus.gov.br/DATASUS/index.php?area=0203 (2020).

52. Brasil. Boletins epidemiologicos. Ministerio da Saude https://www.saude.gov.br/boletins-epidemiologicos (2020).

53. Ten Bosch, Q. A. et al. Contributions from the silent majority dominate dengue virus transmission. PLoS Pathog. 14, e1006965 (2018).

54. Imai, N., Dorigatti, I., Cauchemez, S. & Ferguson, N. M. Estimating Dengue Transmission Intensity from Case-Notification Data from Multiple Countries. PLoS Negl. Trop. Dis. 10, e0004833 (2016).

55. CDC. CDC DENV-1-4 Real-Time RT-PCR Assay. Centers for Disease Control and Prevention https://www.cdc.gov/dengue/resources/rt_pcr/cdcpackageinsert.pdf (2020).

56. Greninger, A. L. et al. Rapid metagenomic identification of viral pathogens in clinical samples by real-time nanopore sequencing analysis. Genome Med. 7, 99 (2015).

57. Greninger, A. L. et al. A metagenomic analysis of pandemic influenza A (2009 H1N1) infection in patients from North America. PLoS One 5, e13381 (2010).

58. Quick, J. et al. Multiplex PCR method for MinION and Illumina sequencing of Zika and other virus genomes directly from clinical samples. Nat. Protoc. 12, 1261–1276 (2017).

59. Grubaugh, N. D. et al. An amplicon-based sequencing framework for accurately measuring intrahost virus diversity using PrimalSeq and iVar. Genome Biol. 20, 8 (2019).

60. Bolger, A. M., Lohse, M. & Usadel, B. Trimmomatic: a flexible trimmer for Illumina sequence data. Bioinformatics 30, 2114–2120 (2014).

61. Li, H. Aligning sequence reads, clone sequences and assembly contigs with BWA-MEM. *arXiv [q-bio.GN]* (2013).

62. Li, H. et al. The Sequence Alignment/Map format and SAMtools. Bioinformatics 25, 2078–2079 (2009).

63. Kearse, M. et al. Geneious Basic: an integrated and extendable desktop software platform for the organization and analysis of sequence data. Bioinformatics 28, 1647–1649 (2012).

64. Katoh, K. & Standley, D. M. MAFFT multiple sequence alignment software version 7: improvements in performance and usability. Mol. Biol. Evol. 30, 772–780 (2013).

65. Nguyen, L.-T., Schmidt, H. A., von Haeseler, A. & Minh, B. Q. IQ-TREE: a fast and effective stochastic algorithm for estimating maximum-likelihood phylogenies. Mol. Biol. Evol. 32, 268–274 (2015).

66. Rambaut, A., Lam, T. T., Carvalho, L. M. & Pybus, O. G. Exploring the temporal structure of heterochronous sequences using TempEst (formerly Path-O-Gen). Virus Evolution vol. 2 vew007 (2016).

67. Suchard, M. A. et al. Bayesian phylogenetic and phylodynamic data integration using BEAST 1.10. Virus Evol 4, vey016 (2018).

68. Hill, V. & Baele, G. Bayesian estimation of past population dynamics in BEAST 1.10 using the Skygrid coalescent model. Mol. Biol. Evol. (2019) doi:10.1093/molbev/msz172.

69. Lemey, P., Rambaut, A., Drummond, A. J. & Suchard, M. A. Bayesian phylogeography finds its roots. PLoS Comput. Biol. 5, e1000520 (2009).

70. Ayres, D. L. et al. BEAGLE 3: Improved Performance, Scaling, and Usability for a High-Performance Computing Library for Statistical Phylogenetics. Syst. Biol. 68, 1052–1061 (2019).

71. Rambaut, A., Drummond, A. J., Xie, D., Baele, G. & Suchard, M. A. Posterior Summarization in Bayesian Phylogenetics Using Tracer 1.7. Syst. Biol. 67, 901–904 (2018).

72. Rambaut, A. FigTree v1.4.4. (2018).

73. Dudas, G. baltic: the Backronymed Adaptable Lightweight Tree Import Code. (2016).

74. Lemey, P., Rambaut, A., Welch, J. J. & Suchard, M. A. Phylogeography takes a relaxed random walk in continuous space and time. Mol. Biol. Evol. 27, 1877–1885 (2010).

75. Dellicour, S., Rose, R., Faria, N. R., Lemey, P. & Pybus, O. G. SERAPHIM: studying environmental rasters and phylogenetically informed movements. Bioinformatics 32, 3204–3206 (2016).

76. Perkins, T. A. et al. An agent-based model of dengue virus transmission shows how uncertainty about breakthrough infections influences vaccination impact projections. PLoS Comput. Biol. 15, e1006710 (2019).

77. Flasche, S. et al. The long-term safety, public health impact, and cost-effectiveness of routine vaccination with a recombinant, live-attenuated dengue vaccine (Dengvaxia): a model comparison study. PLoS Med. 13, e1002181 (2016).

78. Lourenμo, J. et al. Epidemiological and ecological determinants of Zika virus transmission in an urban setting. eLife vol. 6 (2017).

79. Ferguson, N. M. et al. EPIDEMIOLOGY. Countering the Zika epidemic in Latin America. Science 353, 353–354 (2016).

80. Lessler, J. et al. Times to key events in Zika virus infection and implications for blood donation: a systematic review. Bull. World Health Organ. 94, 841–849 (2016).

81. Life expectancy at birth, total (years) - Brazil | Data. https://data.worldbank.org/indicator/SP.DYN.LE00.IN?locations=BR.

82. Yasuno, M. & Tonn, R. J. A study of biting habits of Aedes aegypti in Bangkok, Thailand. Bull. World Health Organ. 43, 319–325 (1970).

83. Trpis, M. & Hausermann, W. Dispersal and other population parameters of Aedes aegypti in an African village and their possible significance in epidemiology of vector-borne diseases. Am. J. Trop. Med. Hyg. 35, 1263–1279 (1986).

84. Li, M. I., Wong, P. S. J., Ng, L. C. & Tan, C. H. Oral susceptibility of Singapore Aedes (Stegomyia) aegypti (Linnaeus) to Zika virus. PLoS Negl. Trop. Dis. 6, e1792 (2012).

85. Wong, P.-S. J., Li, M.-Z. I., Chong, C.-S., Ng, L.-C. & Tan, C.-H. Aedes (Stegomyia) albopictus (Skuse): a potential vector of Zika virus in Singapore. PLoS Negl. Trop. Dis. 7, e2348 (2013).

86. Trpis, M., Häusermann, W. & Craig, G. B. Estimates of Population Size, Dispersal, and Longevity of Domestic Aedes aegypti aegypti (Diptera: Culicidae) by Mark-Release-Recapture in the Village of Shauri Moyo in Eastern Kenya. Journal of Medical Entomology vol. 32 27–33 (1995).

87. Hugo, L. E. et al. Adult survivorship of the dengue mosquito Aedes aegypti varies seasonally in central Vietnam. PLoS Negl. Trop. Dis. 8, e2669 (2014).

